# Characterizing genetic profiles for high triglyceride levels in U.S. patients of African ancestry

**DOI:** 10.1101/2024.03.11.24304107

**Authors:** Lan Jiang, Srushti Gangireddy, Alyson L. Dickson, Yi Xin, Chao Yan, Vivian Kawai, Nancy J. Cox, MacRae F. Linton, Wei-Qi Wei, C. Michael Stein, QiPing Feng

## Abstract

Hypertriglyceridemia (HTG) is a common cardiovascular risk factor characterized by elevated circulating triglyceride (TG) levels. Researchers have assessed the genetic factors that influence HTG in studies focused predominantly on individuals of European ancestry (EA). However, relatively little is known about the contribution of genetic variation to HTG in people of AA, potentially constraining research and treatment opportunities; the lipid profile for African ancestry (AA) populations differs from that of EA populations—which may be partially attributable to genetics. Our objective was to characterize genetic profiles among individuals of AA with mild-to-moderate HTG and severe HTG versus those with normal TGs by leveraging whole genome sequencing (WGS) data and longitudinal electronic health records (EHRs) available in the All of Us (AoU) program. We compared the enrichment of functional variants within five canonical TG metabolism genes, an AA-specific polygenic risk score for TGs, and frequencies of 145 known potentially causal TG variants between patients with HTG and normal TG among a cohort of AA patients (N=15,373). Those with mild-to-moderate HTG (N=342) and severe HTG (N≤20) were more likely to carry *APOA5* p.S19W (OR=1.94, 95% CI [1.48-2.54], p=1.63×10^-6^ and OR=3.65, 95% CI [1.22-10.93], p=0.02, respectively) than those with normal TG. They were also more likely to have an elevated (top 10%) PRS, elevated carriage of potentially causal variant alleles, and carry any genetic risk factor. Alternative definitions of HTG yielded comparable results. In conclusion, individuals of AA with HTG were enriched for genetic risk factors compared to individuals with normal TGs.

## INTRODUCTION

Hypertriglyceridemia (HTG) is a type of hyperlipidemia characterized by elevated circulating triglyceride (TG) levels. Increased TGs are a risk factor for cardiovascular disease, and very high TG levels are also associated with an increased risk of other diseases (e.g., acute pancreatitis).(1–4) HTG is a common condition; an estimated 56.9 million Americans (25.9% of adults ≥20 years old) have elevated TG levels (>150 mg/dL),(5) although very high HTG (>500 mg/dL) is less frequent (1.1% of adults).(6) While the levels used to define HTG vary, genetic studies often define TG concentrations >10 mmol/L or 885 mg/dL as severe HTG.(7–10)

Genetic variation can play a significant role in determining TG levels, particularly among individuals with severe HTG; early sequencing studies identified mutations in five genes canonically associated with TG metabolism (i.e. *LPL, APOA5, APOC2, GPIHBP1,* and *LMF1)* resulting in monogenic HTG.(11) However, monogenic variants explain only a portion of HTG genetics; among patients of European ancestry (EA), only 15.5% of patients with severe HTG and 9% of those with mild-to-moderate HTG have a monogenic determinant of the disease.(8, 10) Moreover, there is also a substantial polygenic contribution to HTG.(8, 10) Despite these advances in understanding genetic contributions to HTG, genetic profiling of HTG in non-EA populations remains limited, hindering our knowledge and potentially constraining research and treatment opportunities, since lipid profiles can vary significantly by ancestry. While genetic studies on inter-individual TG variability have begun to include ancestries beyond those of European patients,(9, 12) relatively little is known about the contribution of genetic variation to high TG levels in people of African ancestry (AA).

Average triglycerides levels are lower in populations of AA than those of EA,(6) and proportionately, fewer individuals of AA than those of EA have very high TG levels.(6, 13–17) While limited, a few studies have examined the relationship between genetics and TG levels in AA individuals. A large-scale GWAS identified different sets of genetic determinants for inter-individual variation of TG levels in people of EA versus those of AA.(18) Further, a polygenic risk score (PRS) derived from AA populations in these studies out-performed a PRS derived from a EA population when tested in external populations of AA.(19) Although these studies are helpful for demonstrating the differential genetic factors between EA and AA populations, they focused largely on individuals with normal or mildly elevated TG levels. There is little information about severe HTG in people of AA; addressing this dearth of information could help facilitate more accurate diagnoses and elucidate future treatment strategies.

Leveraging whole genome sequencing (WGS) data and longitudinal electronic health records (EHRs) available in the All of Us Research Program (AoU), we set out to characterize genetic profiles—including both rare and common variants—among individuals of AA with normal TGs, mild-to-moderate HTG, and severe HTG. Specifically, in these groups, we compared (1) the enrichment of rare and common likely functional variants within the five canonical TG metabolism genes;(11) (2) the PRS for TGs from an AA-specific GWAS study;(19) and (3) the frequencies of 145 potentially causal TG variants in the large Global Lipids Genetics Consortium (GLGC) TG GWAS.(12)

## METHODS

### Data source

Data for the cohort were obtained from the AoU Curated Data Depository (Controlled Tier version 7, released April 20, 2023(20)); AoU is an NIH project that aims to build the most diverse cohort in history. Approximately 45% of participants are racial and ethnic minorities, and over 80% are underrepresented in biomedical research.(21) AoU data include EHRs, genetic information, survey results, and mobile device data as of July 1, 2022 (the cutoff for the version 7 release). All data acquisition, phenotyping, and analyses were completed in the AoU Researcher Workbench, a cloud-based working environment. In accordance with AoU policy, we report categories with fewer than 20 individuals as N≤20 and associated percentages within 5% ranges (e.g., 5-10%, 10-15%); for subgroups with N≤20, but with total N disclosed for the larger category, we report counts as N≤20 and percentages as less than or equal to the percentage represented by 20 individuals (e.g., ≤50%).

### Cohort and demographics

We identified a cohort of adults (i.e., ≥18 years old) who (1) were genetically determined as having predominantly AA,(22) (2) had at least one TG measurement, and (3) had WGS data available in AoU. Although African ancestry and self-reported Black race are largely consistent in AoU (approximately 99% consistency in sampling),(22) to limit the possibility of conflating genetic factors and those associated with social identity, we excluded individuals with a self-reported race other than “Black.” Additionally, we excluded individuals who had a history of (1) metastatic cancer, (2) hospice, (3) feeding tube use, or (4) severe malnutrition based on International Classification of Diseases, Ninth Revision, Clinical Modification (ICD9CM) and International Classification of Diseases, Tenth Revision, Clinical Modification (ICD10CM) diagnosis and procedure codes, Systemized Nomenclature of Medicine (SNOMED) concepts, Healthcare Common Procedure Coding System (HCPCS) codes, and Current Procedural Terminology (CPT) codes (**Supplemental Table S1**). Along with TG measures (described below), we collected gender, age at end of data availability (July 1, 2022), and median BMI from the EHR.

### TG measures and categories

We calculated median TG levels for cohort members from their available TG measures in the AoU EHR. We then extracted lipid-lowering medications for each participant and adjusted median TG levels accordingly (**Supplemental Table S2**).(23–25) Using these adjusted median TG levels (measured TGs), we grouped AA patients into three non-overlapping categories, following previous studies among EA patients:(8–10) (1) severe HTG—patients with median TG levels greater than 885 mg/dL (10 mmol/L); (2) mild-to-moderate HTG—patients with median TG levels between 300 mg/dL and 885 mg/dL; and (3) normal TGs (control)—normolipidemic patients defined as individuals who were not receiving lipid-lowering medications and whose median TG levels fell within the 25^th^-75^th^ percentiles of this cohort (i.e., patients in the 25^th^-75^th^ percentiles who received lipid-lowering medications were excluded because they had the potential to be HTG patients that responded well to treatment, which could obscure genetic differences between the categories, even after adjustment). Given that standards for high TG vary globally and that the prevailing standards have been developed among patient populations with predominantly European ancestry, we conducted sensitivity analyses using two alternative high TG definitions to provide additional nuance to the findings: (1) American Heart Association (AHA) severe HTG (accounting for the study’s setting in the U.S. by using an AHA categorization of severe HTG [>500 mg/dl]), and (2) participants with the top 1% of TG levels (to capture individuals of AA with unusually high TGs for this population group) (**Supplemental Methods**).

### Annotation and analysis of rare and common variants in canonical TG metabolism genes

For analysis of rare variants, we assessed all variants within the canonical TG metabolism gene regions (i.e., Lipoprotein Lipase [*LPL*], chr8:19939253-19967259; Glycosylphosphatidylinositol Anchored High Density Lipoprotein Binding Protein 1 [*GPIHBP1*], chr8:143213218-143217170; Apolipoprotein A5 [*APOA5*], chr11:116789373-116792420; Lipase Maturation Factor 1 [*LMF1*], chr16:853634-970984; and Apolipoprotein C2 [*APOC2*], chr19:44946051-44949565)(11) available in the AoU WGS.(22, 26) All variants were identified and grouped into rare variants (defined as those with minor allele [MAF] frequencies ≤1% in the cohort) and common variants (defined as MAF greater than 1%). We assessed biallelic (homozygous or compound heterozygous) and heterozygous carriage for rare variants collectively and common variants individually; rare and common variants were analyzed separately.

For each variant identified, we used *in silico* prediction algorithms to identify those that were potentially functional, following standards comparable to those detailed in previous publications.(8–10) Specifically, variants were required to have 1) a Combined Annotation Dependent Depletion (CADD) PHRED-scaled score >10, and 2) a prediction of deleterious or damaging function by at least one additional prediction tool—Polymorphism Phenotyping version 2, Sorting Intolerant From Tolerant, or MutationTaster (with a score >0.5)—when classification was available. To provide additional information for those variants identified as potentially functional using this methodology, we also queried the ClinVar database (27, 28) and reported the available ClinVar classifications for genetic variants that were identified as potentially functional in the current study.

### Polygenic risk score for elevated triglyceride (TG) levels

We applied a weighted polygenic risk score for TG levels derived from patients of AA in the Million Veteran Program (MVP);(29) researchers tested multiple models and identified the PRS that explained the highest proportion of variance (R^2^) in TG for MVP patients of African ancestry (available on the PGS catalog, PGS ID accession: PGP000313, **Supplemental Table S3**).(30) Specifically, for our cohort, based on this PRS, we multiplied the number of effect alleles at each locus in the PRS by its associated beta coefficient and then summed these totals across all genetic variants in the PRS (N=285). Then, among normolipidemic individuals (i.e., individuals in the normal category), we determined the minimum score needed to be in the top 10% of the normal category. Finally, we categorized patients in any category with a PRS above that minimum score as having a high polygenic risk score—“top 10% PRS.”

### Potentially causal variants

The GLGC has performed GWAS for lipid-level variation, identifying variants that were potentially causal. Specifically, to account for the potential omission of causal variants given the historical disproportion of EA individuals in GWAS for lipid-level variation, the GLGC conducted broad multi-ancestry GWAS among 5 ancestral groups: admixed African or African, East Asian, European, Hispanic, and South Asian.(12) This analysis identified 145 potentially causal variants for TGs (i.e., >90% posterior probability of being the causal variant at a locus) in ancestry groups and/or multi-ancestry meta-analysis (**Supplemental Table S4**).(12) Given the suggested causality, we sought to determine if these variants accounted for the high TG levels present in some individuals. For these 145 variants, we counted the number of TG-increasing alleles for each individual in our cohort, and those with the highest 10% allele count were termed “top 10% potentially causal TG variants,” and considered to have enriched potentially causal variant carriage.

### Collective Genetic Risk Factors

We identified individuals with any of the genetic risk factors associated with elevated TG levels: carriage of common or rare genetic variants in 5 canonical gene regions; top 10% PRS; or top 10% potentially causal TG variants. These individuals were considered to have elevated overall genetic risk.

### Statistical Analysis

We present cohort characteristics as numbers and percentages for binary variables and medians and IQRs for continuous variables.

For each of the genetic risk factors, given sufficient carriage, we first assessed associations between the factor and measured TGs to confirm a relationship using the entire AA AoU cohort (N=15,373). To test associations between measured TGs and categorical genetic risk factors (i.e., variants in 5 canonical TG genes), we used linear regression with an additive model in PLINK.(31) To assess the relationship between measured TGs and continuous measures of genetic risk factors (i.e., TG PRS and count of potentially causal TG variant alleles), we used Spearman correlations in R.

Then, we compared carriage of genetic risk across TG categories to determine if individuals with HTG were enriched for these factors. For each of the individual genetic risk factors and collective genetic risk, we assessed the odds of carriage in the primary (severe and mild-to-moderate) and sensitivity (AHA severe and top 1%) HTG categories versus the normal TG category. We used Chi-square tests for comparisons with each cell N≥5 and Fisher exact tests for the comparisons with at least one cell N<5).(32–34) Statistical analyses were conducted using R in the AoU Researcher Workbench.

## RESULTS

We identified 15,373 individuals of AA with TG measurements and WGS data available in AoU; of these, 67% were female (N=10,293), with a median [IQR] age of 59 [48-67], and a median [IQR] BMI of 32.00 [27.10-37.99]. Adjusted median measured TGs were 108.40 mg/dL [79.40-149.40] for the cohort. Within the cohort, fewer than 20 individuals (≤0.13%) were in the severe HTG category, and 342 (2.22%) were in the mild-to-moderate HTG category (**Table 1**).

**Table 1.** Cohort characteristics – primary analysis categories.

|  |  | **Severe HTG**<br>(>885 mg/dL) | **Mild-to-moderate HTG**<br>(300-885 mg/dL) | **Normal TG**<br>(79.40-149.40 mg/dL) |
| --- | --- | --- | --- | --- |
| N |  | ≤20 | 342 | 3532 |
| Age (years), median [IQR] |  | 50.5 [40.25-64] | 58 [51-65] | 54 [43-63] |
| Gender, N (%) | Female | ≤20 (70-75%) | 178 (52.0) | 2431 (68.8) |
|  | Male | ≤20 (25-30%) | 164 (48.0) | 1101 (31.2) |
| BMI, median [IQR] |  | 31.89 [27.71-41.01] | 32.30 [28.21-37.25] | 32.47 [27.1-38.6] |
| Lipid-lowering medication, N (%) |  | ≤20 (60-65%) | 273 (79.8) | - |
| Adjusted triglycerides (mg/dL), median [IQR] |  | 1579.0<br>[970.7 -1881.2] | 353.4<br>[321.6-421.3] | 102.5<br>[90.0-120.0] |

IQR=interquartile range; HTG = Hypertriglyceridemia; TG = triglyceride; BMI = body mass index

Normolipidemic individuals with measured TGs between the 25^th^ and 75^th^ percentiles (79.40 and 149.40 mg/dL, respectively) who did not receive lipid-lowering medications were included in the normal TG category (N=3532). Individuals in the severe HTG group were more likely to be younger than individuals in the mild-to-moderate HTG and normal groups, while there was a greater proportion of men in the mild-to-moderate group compared to the others (**Table 1**).

### Canonical TG genes—rare and common variants

In total, from the 5 canonical TG metabolism genes, we identified 239 likely functional genetic variants (**Supplemental Table S5**), including 59 in *LPL*, 17 in *GPIHBP1*, 47 in *APOA5*, 103 in *LMF1*, and 13 in *APOC2*. Of these, 231 were rare genetic variants (i.e., MAF≤1%), and 8 were common variants (i.e., MAF>1%). In total, 40 of these likely functional variants have been reported previously (including 33 rare and 7 common genetic variants) from EA individuals with severe or mild-to-moderate HTG (**Supplemental Table S5**). Additionally, for the 19 variants in these regions identified as pathogenic/likely pathogenic in the ClinVar database, 7 overlapped with our *in silico* prediction (**Supplemental Table S5**), while the remaining 12 variants had no carriage among patients in our HTG groups.

#### 1) Rare genetic variants within canonical TG gene regions

Carriage of biallelic rare variants (either homozygous or compound heterozygous) in the canonical TG metabolism genes was extremely rare in this population; indeed, there were no such carriers among individuals in the severe HTG or mild-to-moderate HTG categories. As such, we did not assess associations with measured TG levels, and we used heterozygous carriage for all additional analyses.

Comparing such carriage across the analysis categories, 20-25% (N≤20) of the severe HTG group and 12.6% (N=43) of the mild-to-moderate HTG group were heterozygous carriers of rare functional variants versus 10.8% (N=383) of the normal TG group (**Figure 1**). There was a trend for individuals in the severe HTG (OR=2.24; 95% CI [0.40-8.54]; p=0.19) and the mild-to-moderate HTG (OR=1.18; 95% CI [0.84-1.66]; p=0.33) groups to be more likely to carry a functional rare variant compared to the normal TG group (**Figure 2, Supplemental Table S6**).

**Figure 1.**
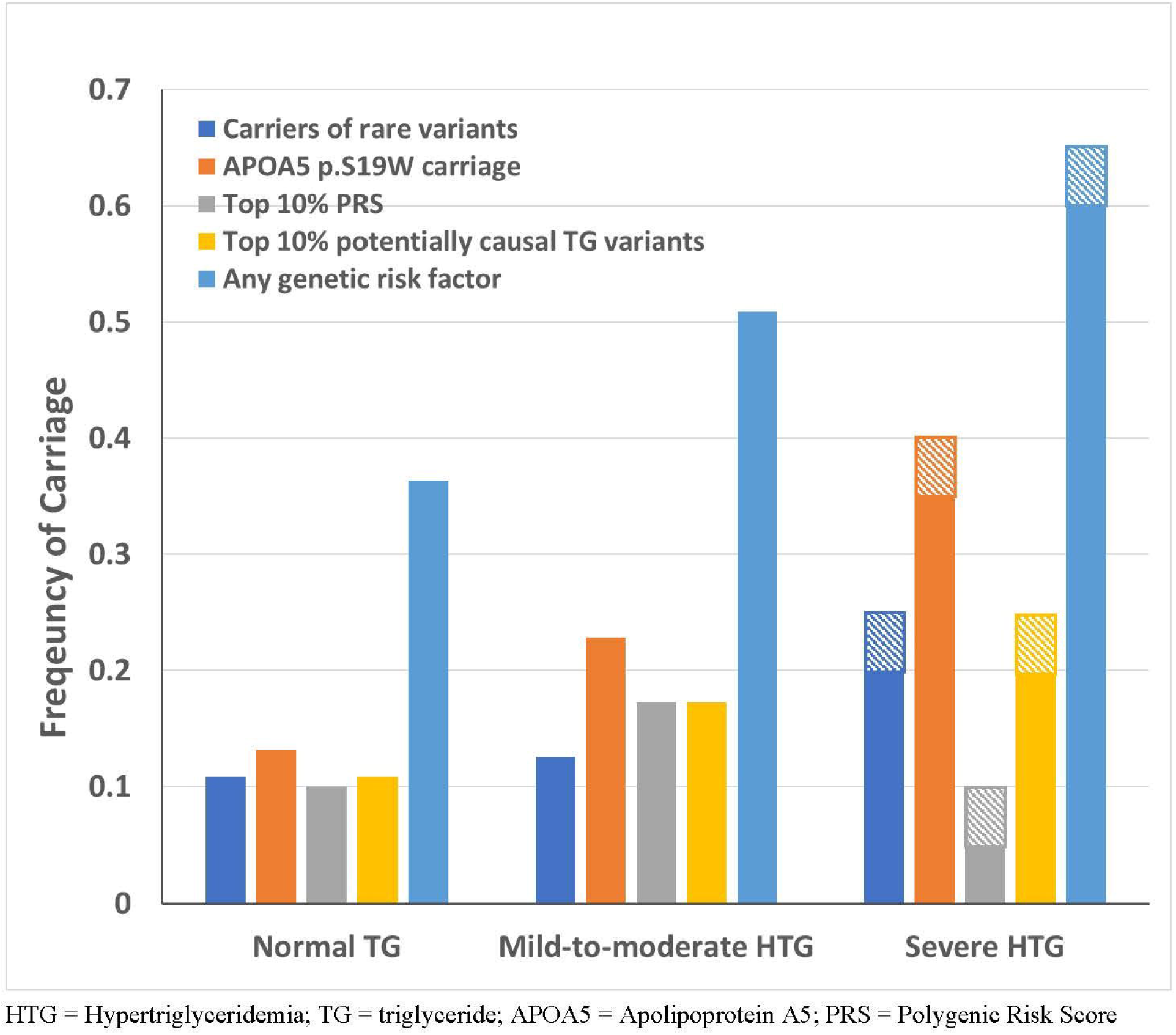
Frequencies of genetic factors in different TG categories (severe HTG, mild-to-moderate HTG, and normal TG) in individuals of African ancestry. *In accordance with AoU policy, percentages for categories with N≤20 are reported in a 5% range (represented by striped bars). HTG = Hypertriglyceridemia; TG = triglyceride; APOA5 = Apolipoprotein A5; PRS = Polygenic Risk Score

**Figure 2.**
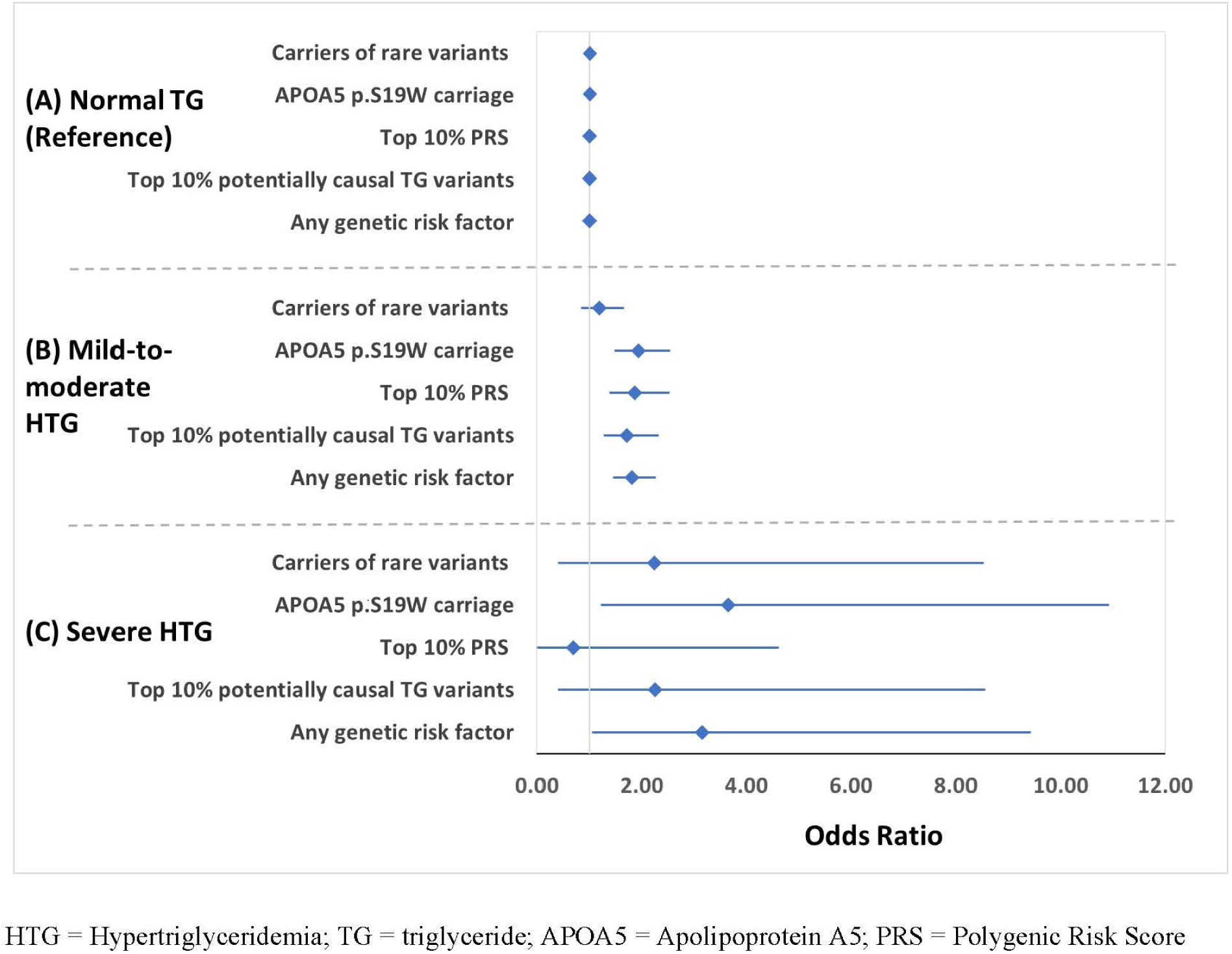
Forest plots for genetic risk factors for elevated TG levels (comparing severe HTG and mild-to-moderate HTG to the normal TGs category). HTG = Hypertriglyceridemia; TG = triglyceride; APOA5 = Apolipoprotein A5; PRS = Polygenic Risk Score

#### 2) Common genetic variants within canonical TG gene regions

The 8 common possible functional variants included 1 variant in *GPIHBP1*, 2 in *APOA5*, and 5 in *LMF1*. When testing the associations between these eight variants and measured TG levels for the cohort, we found that the 5 variants within *LMF1* showed no association with TG levels, and the minor allele of 2 variants (rs78367243 in *GPIHBP1* and rs34282181 in *APOA5*) were associated with lower TG levels (**Supplemental Table S7**), consistent with a recent meta-analysis.(35, 36) Only the minor allele of rs3135506 in *APOA5* was significantly associated in the expected direction with an increase in median measured TG levels (p=1.15×10^-25^), albeit with moderate effect size (β=0.12). The variant has also been associated with TG levels in EA individuals and other ancestries.(37) As such, we only include rs3135506 (*APOA5* p.S19W) in the following comparisons for common genetic variants.

Among the categories, 35-40% (N≤20) and 22.8% (N=78) of the severe HTG and the mild-to-moderate HTG categories were carriers of *APOA5* p.S19W, respectively. In contrast, only 13.2% (N=467) of the normal TG group carried the variant. Individuals in the severe HTG and the mild-to-moderate HTG categories were 3.65 times (95% CI [1.22-10.93]; p=0.02) and 1.94 times (95% CI [1.48-2.54]; p=1.63×10^-6^) more likely to have *APOA5* p.S19W than the normal TG category, respectively (**Figure 2, Supplemental Table S6**).

### AA-derived TG PRS

All 285 SNPs comprising the TG PRS derived from a cohort of individuals of African ancestry were available for analyses.(19, 30) The TG PRS was significantly correlated with measured TG levels (r=0.12, p<2.2×10^-16^, N=15,373) and its distribution in each category is shown in **Supplemental Table S8**. Individuals with an elevated TG PRS (defined as top 10% in the normal TG group) represented 5-10% (N≤20), 17.3% (N=59), and 10.0% (N=354) of the severe HTG, mild-to-moderate HTG, and normal TG categories, respectively (**Figure 1**).

Although we found no increased odds of elevated TG PRS in the severe HTG category relative to the normal TG category (OR=0.69, 95% CI [0.02-4.62]; p=1.00), individuals in the mild-to-moderate HTG category were 1.87 times (95% CI [1.38-2.53]; p=4.5×10^-5^) more likely to carry a high TG PRS compared to those in the normal TG category (**Figure 2, Supplemental Table S6**).

### Accumulation of potentially causal TG variants

Of the 145 potentially causal TG variants previously identified by the GCLC multi-ancestry GWAS, 140 were available for our cohort (including one variant also assessed in the 5 canonical TG gene regions, **Supplemental Table S4**). From these 140 available variants, we calculated the number of potentially causal TG variant alleles (maximum possible alleles, N=280) carried by each cohort member. The number of alleles (median [IQR], 132 [128, 136]) significantly correlated with measured TG levels in the cohort (r=0.06, p=2.71×10^-13^, N=15,373). The carriage of potentially causal TG variant alleles in each group are shown in **Supplemental Table S9**.

Individuals with an elevated carriage of potentially causal TG variants (top 10 % in our cohort [i.e., N >140 alleles]) represented 20-25% (N≤20), 17.3% (N=59), and 10.8% (N=382) of the severe HTG, mild-to-moderate HTG, and normal TG groups, respectively (**Figure 1**). Compared to the normal TG control group, individuals with severe or mild-to-moderate HTG were 2.25 times (95% CI [0.40-8.56]; p=0.19) and 1.72 times (95% CI [1.27-2.32]; p=4.0×10^-4^) more likely to have elevated carriage of potentially causal TG variants, respectively (**Figure 2, Supplemental Table S6**).

### Collective Genetic Risk Factors

Finally, we compared the likelihood of having any genetic risk factors for elevated TG levels in each TG category. Individuals with at least one genetic risk factor represented 60-65% (N≤20) and 50.9% (N=174) of the severe HTG and mild-to-moderate HTG categories, respectively; in comparison, individuals with at least one genetic risk factor represented only 36.4% (N=1284) of the normal TG category (**Figure 1**). Relative to the normal TG category, the severe HTG and the mild-to-moderate HTG categories were 3.15 times (95% CI [1.05-9.42], p-value=0.04) and 1.81 times more likely (95% CI [1.45-2.27], p=1.65×10^-7^) to have at least one genetic risk factor related to high TGs, respectively (**Figure 2, Supplemental Table S6**).

### Sensitivity analyses

Sensitivity analyses in two additional high TG groups (i.e., AHA severe HTG and Top 1% TG) were consistent with the main analyses (**Supplemental Results, Supplemental Figures S1 & S2,** and **Supplemental Tables S10 & S11**).

## DISCUSSION

We completed the first assessment of genetic profiles for HTG in individuals with predominantly African ancestry. We observed enrichment of genetic factors (i.e., functional genetic variants in 5 canonical TG genes and potentially causal TG genetic variants) in the severe HTG and mild-to-moderate HTG categories compared to individuals in the normal TG category. These observations were further confirmed in analyses that used two additional definitions of HTG.

We identified one common likely functional variant in *APOA5* (rs3135506, p.S19W) that was significantly associated with elevated TGs. This variant has been reported as associated with increased TGs in EA and Hispanic populations, with minor allele frequencies (MAFs) of 6.37% and 15.27%, respectively;(9) we observed an MAF of 6.29% in this cohort of individuals with AA. Notably, carriers of *APOA5* p.S19W were significantly overrepresented in the severe HTG category compared to the normal TG category (35-40% versus 13.2%, p=0.02). Our observation confirmed that *APOA5* p.S19W impacts TG metabolism in the AA population; however, that effect may be moderate, as indicated by the effect size (β=0.12). Moreover, we did not find other common likely functional genetic variants in severe or mild-to-moderate HTG; notably, 2 SNPs (rs78367243 in *GPIHBP1* and rs34282181 in *APOA5*) were associated with lower TG levels, consistent with previous meta-analyses in cohorts of EA and East Asian ancestry.(35, 36) Overall, these results suggest that common variants in these canonical TG-associated genes have limited effects on HTG in AA individuals.

Populations with African ancestry generally include more genetic variation compared to other ancestries,(38–42) including rare functional variants. Indeed, we identified 231 rare likely functional variants (out of 239 in total) from AA individuals in AoU. Of these, only 40 have been reported from EAs with severe or mild-to-moderate HTG. The MAFs are comparable between AA cohort (AoU) and 1000 genome project African ancestry (AFR) (**Supplemental Table S5**). In our analysis, we observed 20-25% of the severe HTG AA category were heterozygous for at least one rare variant, compared to 12.5-15% reported previously in EA patients with severe HTG (combined biallelic and heterozygous carriers).(8, 9) Similarly, in the mild-to-moderate HTG category (300-885 mg/dL), 12.6% of AA individuals carried heterozygous rare variants, higher than the 9% of EA patients reported for the same category.(10) We note that the use of whole genome sequencing data from a large number of patients (N=15,373) likely contributed to the increased number of rare functional variants observed in the current study—most previous studies conducted exome sequencing in a smaller number of EA patients. It is possible that large studies using whole genome sequencing could yield higher rates of heterozygous rare variant carriage among EA populations.

Notably, despite the enrichment of rare variants in AAs, we did not find any biallelic carriers (homozygous or compound homozygous) of likely functional variants in AAs with HTG (severe or mild-to-moderate). Previous studies have shown that individuals of AA have lower average TGs and proportionally fewer cases of HTG.(17) The lack of biallelic carriers of functional variants in critical TG genes (e.g., the five canonical genes tested in this study) might explain, at least partially, the favorable TG profile in AA individuals.

In the assessment of the AA-derived TG PRS, we did not find an enrichment of PRS (top 10%) in the severe HTG AAs. While there was an enrichment of elevated PRS in the mild-to-moderate HTG (17.3%) compared to normal TG controls (10.0%), the prevalence of elevated PRS was lower than previously reported from EA populations using a PRS derived from a EA population (i.e., in a previous report from EAs, 26.9% of the individuals with mild-to-moderate HTG with an elevated EA-derived PRS).(10) Several factors may limit the interpretation of these differences: (1) our severe HTG group was small; and 2) the PRS were derived from ancestry-specific cohorts and differed in the number of SNPs included (N=285 for AA; N=16 for EA).(8, 19, 43) Despite these factors, the low prevalence of elevated PRS in the severe HTG group indicates that the genetic risk defined by the PRS may play a smaller role in HTG for individuals of AA compared to EA.

The carriage of 145 potentially causal variants has not been tested previously among AA individuals. We found enrichment of potentially causal variants in all HTG categories (both primary and sensitivity analyses) compared to the normal TG categories.

Considering all genetic factors, we observed a clear enrichment in AA individuals with HTG— more than half of the individuals any HTG category (primary or sensitivity) had at least one contributing genetic factor, while only 36.4% of normolipidemic AA individuals had one. However, it is notable that approximately 35-40% of AA individuals with severe HTG and half of those with mild-to-moderate HTG did not have identifiable genetic factors. This finding raises several possibilities. First, the genetics of HTG are not fully understood. While large GWAS are available for TGs, most studies include patients with normal TG levels, and patients with HTG are rare or even absent. Second, although existing GWAS are powerful for identifying common variants, statistical power is more limited for detecting rare functional variants, such as those in the canonical TG genes. Third, there have been fewer studies focused on AA populations and almost no genetic analysis on AA individuals with HTG. Indeed, even the definitions of HTG are derived from populations that are predominantly EA. Fourth, for those variants involved in TG metabolism, most genetic analyses have assumed a linear relationship, which may not be true for the variants with larger effect size. Lastly, comorbidities (e.g., type 2 diabetes), lifestyle (e.g., smoking, alcohol intake, and diet), and secondary factors (e.g., other medications) can also influence measured TGs, but the pervasiveness or lack of granularity associated with these factors limited our ability to incorporate either additional exclusions or adjustments, respectively.

This study offers several strengths. It is the first study focusing on the genetic profiles of AA individuals with HTG. Second, by leveraging the medication information from EHRs, we were able to adjust the TG measurement by known lipid medications, reducing potential misclassification. Third, compared to previous studies that focused on exome sequencing data, we were able to utilize WGS and identify more potential functional variants from the 5 canonical TG gene regions. Fourth, we applied a sophisticated AA-derived TG PRS to our AA cohort. This AA PRS has outperformed EA-derived PRS in cohorts of African ancestry.(19) Fifth, as sensitivity analyses, we assessed two additional HTG categories, including the AHA severe HTG cutoff (>500mg/dl) and the top 1% of measured TGs in the cohort, yielding comparable results.

Our study also has some limitations. First, our severe HTG category was small, limiting our analyses. According to the previous population reports, the prevalence for multifactorial chylomicronemia syndrome (MCS) and familial chylomicronemia syndrome (FCS) are 1:600 and 1-10:1,000,000, respectively.(44) Therefore, the current study cohort (N=15,373) lacked sufficient sample size for meaningful analysis of such conditions among severe HTG cases —in fact, we did not identify any biallelic LOF variant carriers across the four HTG groups in either primary and sensitivity analyses. Likewise, as noted in results, among the 19 variants identified as pathogenic/likely pathogenic in the ClinVar database, 7 overlapped with our *in silico* prediction. However, for the 12 additional potentially functional variants that did not overlap with *in silico* prediction, there was no carriage among individuals in any of our HTG groups.

This observation is consistent with the rarity of many such pathogenic variants and suggests that future studies in larger cohorts are needed, especially for populations of AA. Moreover, this limited sample size also restricted our ability to meaningfully report additional clinical factors for HTG patients, given the data privacy restrictions of AoU. Second, given that the clinical data was restricted primarily to EHR in AoU, for patients with severe HTG, we could not gather additional detailed background and clinical information, as might be collected routinely in a specialty clinical setting or clinical trial. Third, while we focused on AA individuals, we did not have enough power to explore HTG in other non-European ancestries or address the impact of admixture. Fourth, we used *in silico* prediction software to determine functionality of genetic variants in the 5 canonical TG genes. We did not conduct *in vitro* analyses to confirm their function, and future function research is needed. Fifth, due to limited statistical power, we could not test the association of rare variants with measured TGs. Sixth, we did not include lifestyle and secondary factors as covariates in our analyses. The AoU program has started to collect social determinants of health (SDOH) factors using questionnaires, and the data are growing. Future analyses are needed to address the role of these factors in HTG.

In conclusion, we have laid the groundwork for a more complete understanding of the genetic factors associated with elevated TG levels among individuals with AA. As we continue to expand this understanding, such knowledge can contribute to better predicting risk and determining best practices for clinical interventions in the future.

## Supporting information

supplementary methods and results

## Data Availability

All data produced are available online at All of Us (AoU)

## Data and Code Availability Statement

In line with the privacy standards set by the All of Us Research Program, data and code used for this study are available to approved researchers who register for access to the Researcher Workbench platform at https://workbench.researchallofus.org/login. This analysis was run on the All of Us Research Program Controlled Tier Dataset version 7, production release C2022Q4R9.

## Acknowledgements

The All of Us Research Program would not be possible without the partnership of its participants. Additionally, the All of Us Research Program is supported by the National Institutes of Health, Office of the Director: Regional Medical Centers: 1 OT2 OD026549; 1 OT2 OD026554; 1 OT2 OD026557; 1 OT2 OD026556; 1 OT2 OD026550; 1 OT2 OD 026552; 1 OT2 OD026553; 1 OT2 OD026548; 1 OT2 OD026551; 1 OT2 OD026555; IAA #: AOD 16037; Federally Qualified Health Centers: HHSN 263201600085U; Data and Research Center: 5 U2C OD023196; Biobank: 1 U24 OD023121; The Participant Center: U24 OD023176; Participant Technology Systems Center: 1 U24 OD023163; Communications and Engagement: 3 OT2 OD023205; 3 OT2 OD023206; and Community Partners: 1 OT2 OD025277; 3 OT2 OD025315; 1 OT2 OD025337; 1 OT2 OD025276.

## Author Contributions

**Lan Jiang** – Conceptualization, Data curation, Formal analysis, Writing - review & editing; **Srushti Gangireddy** – Data curation, Writing - review & editing; **Alyson L. Dickson** – Data curation, Formal analysis, Writing - Original Draft, Writing - review & editing; **Yi Xin –** Data curation, Writing - review & editing; **Chao Yan –** Data curation, Writing - review & editing; **Vivian Kawai** – Methodology, Writing - review & editing; **Nancy J. Cox** – Conceptualization, Writing - review & editing; **MacRae F. Linton** – Methodology, Writing - review & editing; **Wei-Qi Wei** – Funding acquisition, Methodology, Software, Writing - review & editing; **C. Michael Stein** – Conceptualization, Methodology, Formal analysis, Writing - Original Draft; Writing - review & editing; **QiPing Feng** – Conceptualization, Formal analysis, Funding acquisition, Project administration, Writing - Original Draft, Writing - review & editing

