## supplementary methods and results for "Characterizing genetic profiles for high triglyceride levels in U.S. patients of African ancestry"

**Supplemental Methods**

Sensitivity Analysis Categories

Given that standards for high triglycerides TG) vary globally and that the prevailing standards have been developed among patient populations with predominantly European ancestry (EA), we conducted sensitivity analyses using two alternative high TG definitions to provide additional nuance to the assessment of high TG levels among patients of African ancestry (AA) in the U.S. First, to account for accepted U.S. standards, we designated an American Heart Association (AHA) severe hypertriglyceridemia (HTG) category—individuals with median TG levels greater than 500 mg/dL, as defined by AHA guidelines. Second, to address the possibility that standards developed primarily among patients with EA may not most effectively identify “high” TG levels among individuals with AA, we designated a top 1% TG category—individuals with TG levels greater than 99% of AA patients in our cohort. These two categories (i.e., AHA severe HTG and top 1%) included overlapping individuals with the primary analysis categories of severe HTG and mild-to-moderate HTG. We analyzed each of these two categories separately.

**Supplemental Results**

We first identified 54 individuals in the AHA severe HTG category (i.e., individuals with median TG levels greater than 500 mg/dL); of these, 29 (53.7%) were female and the median age [IQR] was 55.00 [48.00-64.00]). We identified those with median TG level greater than 374.4 mg/dL as the top 1% TG category. Of the 154 individuals in this category, 77 (50.0%) were female and the median age [IQR] was 57.00 [49.00-64.00] (**Supplemental Table S10**).

Canonical TG genes—rare and common variants

1) Rare genetic variants within canonical TG gene regions

Less than 37.5% (N≤20) of the AHA severe HTG category and 18.8% (N=29) of the top 1% TG category carried rare functional variants (**Supplemental Figure S1**). Individuals in the AHA severe HTG and top 1% TG categories were 1.87 times (95% CI [0.93-3.74]; p=0.078) and 1.91 times (95% CI [1.26-2.90]; p=0.002) more likely to carry a rare variant compared to the normal TG group, respectively (**Supplemental Figure S2, Supplemental Table S11**).

2) Common genetic variants within canonical TG gene regions

Less than 37.5% (N≤20) of the AHA severe HTG category and 20.8% (N=32) of the top 1% TG category carried the APOA5 p.S19W variant. Individuals in the AHA severe HTG and top 1% TG categories were 1.88 times (95% CI [0.98-3.59]; p=0.057) and 1.72 times (95% CI [1.15-2.57]; p=0.009) more likely to carry a rare variant compared to the normal TG group, respectively (**Supplemental Table S11**).

AA-derived TG polygenic risk score (PRS)

Elevated (i.e., top 10%) TG PRS were found in ≤37.5% (N≤20) and 16.2% (N=25) of the AHA severe HTG and top 1% TG categories, respectively (**Supplementary Figure 2**). The resulting odds ratios of having elevated TG PRS when compared to individuals in the normal TG category were similar to those of the mild-to-moderate HTG category: 1.56 (95% CI [0.73-3.33]; p=0.25) and 1.74 (95% CI [1.12-2.71]; p=0.014) for the AHA severe HTG and top 1% TG categories, respectively (**Supplementary Figure 2, Supplementary Table 11**).

Accumulation of potentially causal TG variants

Enrichment (i.e., top 10% allele count) for the 145 potentially causal TG variants was identified in ≤37.5% (N≤20) and 16.9% (N=26) of AA individuals in the AHA severe HTG and top 1% TG categories, respectively (**Supplementary Figure 1**). AA individuals in the top AHA severe HTG and 1% TG categories were more likely to be enriched for potentially causal TG variants (OR=2.11, 95% CI [1.08-4.13]; p=0.029 and OR=1.67, 95% CI [1.08-2.59]; p=0.02, respectively) compared to the normal TG category (**Supplementary Figure 2, Supplementary Table 11**).

Collective Genetic Risk Factors

Patients with at least one genetic risk factor represented 57.4% (N=31) and 55.2% (N=85) of the AHA severe HTG and the top 1% TG categories (**Supplementary Figure 1**). When compared to the normal TG category, individuals in the AHA severe HTG and top 1% TG were 2.36 and 2.16 times more likely (95% CI [1.37-4.06], p-value=0.002 and 95% CI [1.56-2.98], p=3.54×10^-6^, respectively) to carry a genetic risk factor (**Supplementary Figure 2, Supplementary Table 11**).
