## Supplementary tables and figures for "Characterizing genetic profiles for high triglyceride levels in U.S. patients of African ancestry"

**Table of Contents**

Supplemental Table S1. ICD Codes for Excluded Conditions.

Supplemental Table S2. Lipid-lowering Medications and Related Triglyceride Adjustments.

Supplemental Table S3. SNPs and Weights of a Polygenic Risk Score for Triglycerides Derived from a Population with African Ancestry.

Supplemental Table S4. Potentially Causal Variants for Triglycerides Identified by the Global Lipids Genetic Consortium.

Supplemental Table S5. Annotation of Likely Functional Variants within 5 Canonical Triglyceride Gene Regions.

Supplemental Table S6. Accumulation of Genetic Risk Factors in Mild-to-moderate HTG and Severe HTG categories, Compared to Normal TG Controls—Primary Analyses.

Supplemental Table S7. Association between Common Variants from 5 Canonical Triglyceride Gene Regions and Log-Transformed Median Measured Triglycerides Adjusted by Gender and Age.

Supplemental Table S8. Polygenic Risk Scores for Triglycerides Derived from a Population with African Ancestry by Category.

Supplemental Table S9. Total Allele Count of Potentially Causal Triglyceride Variants by Category.

Supplemental Table 10. Cohort Characteristics—Sensitivity Analysis Categories (AHA Severe HTG and Top 1% TG).

Supplemental Figure S1. Sensitivity Analyses: Frequencies of Genetic Factors in Different TG Categories (AHA Severe HTG, Mild-to-Moderate HTG, and Normal TG) in Individuals of African Ancestry.

Supplemental Figure S2. Sensitivity Analyses: Forrest Plots for Genetic Risk Factors for Elevated TG Levels in Individuals of African ancestry. (AHA Severe HTG, Mild-to-Moderate HTG, and Normal TG).

Supplemental Table S11. Accumulation of Genetic Risk Factors in AHA Severe HTG, and Top 1% HTG categories, Compared to Normal TG Controls—Sensitivity Analyses.

**Supplemental Table S1. ICD Codes for Excluded Conditions.**

| **Exclusion Condition** | **Code Type** | **Code** |
| --- | --- | --- |
| Malnutrition | ICD10CM | E40, E41, E42, E43, E45, E46, E64, H46.2, P00.4, R64 |
|  | ICD9CM | 260, 261, 262, 263, 263.8, 263.9, 579.3, 760.4, 799.4 |
|  | SNOMED | 2492009, 360549009 |
| Metastatic cancer | CPT | 49255, 69150, 3301F |
|  | HCPCS | G9066, G9069, G9075, G9087, G9088, G9094, G9098, G9103, G9107, G9111, G9838, G9842 |
|  | ICD10CM | C77.0, C77.1, C77.2, C77.3, C77.4, C77.5, C77.8, C77.9, C78, C78.0, C78.00, C78.01, C78.02, C78.1, C78.2, C78.3, C78.30, C78.39, C78.4, C78.5, C78.6, C78.7, C78.8, C78.80, C78.89, C79, C79.0, C79.00, C79.01, C79.02, C79.1, C79.10, C79.11, C79.19, C79.2, C79.3, C79.31, C79.32, C79.4, C79.40, C79.49, C79.5, C79.51, C79.52, C79.6, C79.60, C79.61, C79.62, C79.63, C79.7, C79.70, C79.71, C79.72, C79.8, C79.81, C79.82, C79.89, C79.9, C7B, C7B.0, C7B.00, C7B.01, C7B.02, C7B.03, C7B.04, C7B.09, C7B.1, C7B.8 |
|  | ICD9CM | 196.0, 196.1, 196.2, 196.3, 196.5, 196.6, 196.8, 196.9, 197, 197.0, 197.1, 197.2, 197.3, 197.4, 197.5, 197.6, 197.7, 197.8, 198.0, 198.1, 198.2, 198.3, 198.4, 198.5, 198.6, 198.7, 198.8, 198.81, 198.82, 198.89, 209.7, 209.70, 209.71, 209.72, 209.73, 209.74, 209.75, 209.79 |
|  | SNOMED | 61517002, 94161006, 94180008, 94181007, 94183005, 94186002, 94217008, 94222008, 94225005, 94243009, 94246001, 94264008, 94280003, 94297009, 94298004, 94313005, 94326009, 94339008, 94340005, 94346004, 94347008, 94348003, 94350006, 94351005, 94352003, 94360002, 94365007, 94381002, 94386007, 94391008, 94392001, 94393006, 94394000, 94395004, 94396003, 94397007, 94398002, 94400003, 94408005, 94409002, 94416001, 94441008, 94442001, 94455000, 94474008, 94480000, 94493005, 94515004, 94519005, 94579000, 94580002, 94582005, 94595000, 94600009, 94602001, 94603006, 94614009, 94615005, 94627008, 94628003, 94634005, 94649002, 94654006, 94663008, 94664002, 94678001, 128462008, 193445006, 275266006, 307226002, 314994000, 315241008, 359780007, 359785002, 369523007, 369530001, 402378006, 403906006, 405843009, 443493003, 702392008, 705176003, 712849003, 722671009, 91281000119103, 353561000119103, 353741000119106, 457721000124104, 459381000124106, 459391000124109, 461511000124101, 1082601000112100 |
| Feeding tube | CPT | 42955, 43653, 43760, 43761, 43762, 43763, 43830, 43832, 43870, 44015, 44300, 44372, 44373, 49440, 49441, 49446, 49450, 49451, 49460, 49465, 0647T |
|  | HCPCS | B4034, B4035, B4036, B4085, B4086, B4087, B4088, B4102, B4103, B4104, B4105, B4149, B4150, B4152, B4153, B4154, B4155, B4157, B4164, B4168, B4172, B4176, B4178, B4180, B4185, B4189, B4193, B4197, B4199, B4216, B4220, B4222, B4224, B5000, B5100, B5200, B9002, B9004, B9006, B9998, B9999, E0791 |
|  | ICD10CM | K94.2, K94.20, K94.21, K94.22, K94.23, K94.29, Z43.1, Z93.1 |
|  | ICD10PCS | 0DH50UZ, 0DH53UZ, 0DH54UZ, 0DH57UZ, 0DH58UZ, 0DH60UZ, 0DH63UZ, 0DH64UZ, 0DH67UZ, 0DH68UZ, 0DH80UZ, 0DH83UZ, 0DH84UZ, 0DH87UZ, 0DH88UZ, 0DH90UZ, 0DH93UZ, 0DH94UZ, 0DH97UZ, 0DH98UZ, 0DHA0UZ, 0DHA3UZ, 0DHA4UZ, 0DHA7UZ, 0DHA8UZ, 0DHB0UZ, 0DHB3UZ, 0DHB4UZ, 0DHB7UZ, 0DHB8UZ, 0DP60UZ, 0DP63UZ, 0DP64UZ, 0DP67UZ, 0DP68UZ, 0DW00UZ, 0DW03UZ, 0DW04UZ, 0DW07UZ, 0DW08UZ, 0DW0XUZ, 0DW60UZ, 0DW63UZ, 0DW64UZ, 0DW67UZ, 0DW68UZ, 0DW6XUZ, 0DWD0UZ, 0DWD3UZ, 0DWD4UZ, 0DWD7UZ, 0DWD8UZ, 0DWDXUZ, 0D20XUZ, 3E0G36Z, 3E0G76Z, 3E0G86Z |
|  | ICD9CM | 536.4, 536.40, 536.41, 536.42, 536.49, V44.1, V55.1 |
|  | ICD9Proc | 43.0, 43.1, 43.11, 43.19, 46.3, 46.31, 46.32, 46.39, 97.02 |
|  | SNOMED | 6125005, 61420007 |
| Hospice | CPT | 99377, 99378 |
|  | HCPCS | G0031, G0034, G0048, G0051, G0182, G0299, G0300, G0337, G0493, G0494, G0495, G0496, G2153, G9054, G9433, G9473, G9474, G9475, G9476, G9477, G9478, G9479, G9524, G9687, G9688, G9690, G9691, G9692, G9693, G9694, G9700, G9702, G9707, G9709, G9710, G9713, G9714, G9715, G9718, G9720, G9723, G9725, G9740, G9741, G9758, G9760, G9761, G9768, G9802, G9805, G9809, G9819, G9857, G9858, G9860, G9861, G9988, G9992, G9993, G9994, G9995, G9996, M1022, M1025, M1026, M1059, M1067, M1154, M1159, M1165, M1167, M1186, M1191, Q5003, Q5004, Q5005, Q5006, Q5007, Q5008, Q5010, S0255, S0271, S9126, T2042, T2043, T2044, T2045, T2046 |
|  | ICD10CM | R40.3, Z51.5, Z66 |
|  | ICD9CM | 780.03, V49.86, V66.7 |

CPT = Current Procedural Terminology; HCPCS = Healthcare Common Procedure Coding System; ICD10CM = International Classification of Diseases, Tenth Revision, Clinical Modification diagnosis codes; ICD10PCS = International Classification of Diseases, Tenth Revision, Clinical Modification procedure codes; ICD9CM = International Classification of Diseases, Ninth Revision, Clinical Modification diagnosis codes; ICD9Proc = International Classification of Diseases, Ninth Revision, Clinical Modification procedure codes; SNOMED = Systemized Nomenclature of Medicine concepts

**Supplemental Table S2. Lipid-lowering Medications and Related Triglyceride Adjustments.**

| **Adjustment Type^a^** | **TG Constant**(1, 2) | **Medication Type** | **Search Terms^b^** |
| --- | --- | --- | --- |
| Fibrates | 57.1 | fibrate | bezafibrate, Bezalip, Atromid-s, clofibrate, Antara, Atorva TG, Fenocor, fenofibrate, Fenogal, Fenoglide, Fibricor, Golip, Lipanthyl, Lipantil, Lipidil, Lipofen, Lofibra, Phenofibrate, Procetofen, Supralip, Tricheck, Tricor, Triglide, fenofibric acid, Trilipix, fibric acid, gemfibrozil, Lopid |
|  |  | statin & fibrate | fenofibrate-pravastatin, Pravafenix, Cholib, fenofibrate-simvastatin |
| Prescription omega-3 fatty acids | 45.6 | omega-3-ethyl esters/ icosapent ethyl | Lovaza, Omacor, Vascepa |
| Niacin | 89.4 | niacin | Endur-acin, niacin, Niacin SR, Niacin-50, Niacor, Slo-Niacin, Endur-Amide, niacinamide, Niaspan, nicotinamide, nicotinamide adenine dinucleotide, nicotinamide riboside, Tru Niagen, nicotinic acid, vitamin B3 |
|  |  | statin & niacin | Advicor, lovastatin-niacin, Simcor, simvastatin-niacin |
| Statins | 18.4 | statin | amolodipine-atorvastatin, Caduet, Atorvaliq, atorvastatin, Lipitor, Baycol, cerivastatin, Lipobay, fluvastatin, Lescol, Altocor, Altoprev, lovastatin, Mevacor, Livalo, pitavastatin, Zypitamag, Pravachol, pravastatin, pravastatin-aspirin, Pravigard, Crestor, Ezallor, rosuvastatin, FloLipid, simvastatin, Zocor, Juvisync, sitagliptin-simvastatin |
|  |  | statin & CAI | ezetimibe-atorvastatin, Liptruzet, ezetimibe-rosuvastatin, Roszet, ezetimibe-simvastatin, Vytorin |
| ACLI | 0 | ACLI | Nexletol, Nilemdo, bempedoic acid |
| Bile acid sequestrants | 0 | bile acid sequestrants | Prevalite, cholestyramine, Questran, Welchol, Cholestagel, colesevelam, Lodalis, Colestid, colestipol, Cholestabyl, Lestid |
| CAI | 0 | CAI | Zetia, ezetimibe |
|  |  | CAI & ACLI | Nexlizet, Nustendi, bempedoic acid-ezetimibe |
| PCSK9 inhibitors | 0^c^ | PCSK9 inhibitor/monoclonal antibody | Praluent, alirocumab, Repatha, evolocumab |

^a^ Dual category prescriptions (e.g., statin-fibrate combinations) were assigned to the larger of the two adjustments.

^b^ Prescription entries in the electronic health records were excluded if they included the strings “patch,” “topical,” “mask,” “cream,” “ointment,” “gel,” “shampoo,” “liquid,” “spray,” “emulsion,” “lotion,” “serum,” “hair,” “prenatal,” “skin,” “miracle,” or “dermatology.”

^c^ Although PSCK9 inhibitors can lower triglycerides,(3, 4) no constant for adjustment has been determined; however, given their relatively recent approval, this lack of adjustment affected very few individuals in the cohort (N≤20) and none of the individuals in the HTG groups (for either the primary or sensitivity analyses).

ACLI = ATP Citrate Lyase inhibitors; CAI = Calcium Absorption inhibitors; PSCK9 = Proprotein convertase subtilisin/kexin type 9

**Supplemental Table S3. SNPs and Weights of a Polygenic Risk Score for Triglycerides Derived from a Population with African Ancestry**.(5, 6)

| **rsID** | **chr_name** | **effect_allele** | **other_allele** | **effect_weight** | **hm_source** | **hm_pos** | **hm_inferOtherAllele** |
| --- | --- | --- | --- | --- | --- | --- | --- |
| rs72911746 | 1 | T | C | -0.034127 | ENSEMBL | 54746394 | TRUE |
| rs146524463 | 1 | T | C | -0.0369882 | ENSEMBL | 54917162 | TRUE |
| rs72907616 | 1 | A | C | -0.0228964 | ENSEMBL | 54981312 | TRUE |
| rs115140720 | 1 | A | G | -0.0404033 | ENSEMBL | 54981711 | TRUE |
| rs2249480 | 1 | T | C | 0.01025902 | ENSEMBL | 54990863 | TRUE |
| rs191261701 | 1 | A | G | -0.0606213 | ENSEMBL | 55014918 | TRUE |
| rs2479395 | 1 | T | C | -0.0160994 | ENSEMBL | 55018909 | TRUE |
| rs7525649 | 1 | T | C | 0.00969948 | ENSEMBL | 55033483 | TRUE |
| rs6681159 | 1 | T | C | 0.01784408 | ENSEMBL | 55042209 | TRUE |
| rs7543163 | 1 | T | C | 0.01623986 | ENSEMBL | 55049808 | TRUE |
| rs11806638 | 1 | A | C | -0.0276708 | ENSEMBL | 55052487 | TRUE |
| rs1165287 | 1 | A | G | 0.01980777 | ENSEMBL | 55054539 | TRUE |
| rs28362258 | 1 | T | C | 0.01128212 | ENSEMBL | 55057123 | TRUE |
| rs509504 | 1 | A | G | 0.02440458 | ENSEMBL | 55057360 | TRUE |
| rs28362261 | 1 | G | A | -0.0433872 | ENSEMBL | 55058129 | TRUE |
| rs28362263 | 1 | A | G | -0.0522555 | ENSEMBL | 55058182 | TRUE |
| rs141502002 | 1 | T | C | 0.02435385 | ENSEMBL | 55058549 | TRUE |
| rs505151 | 1 | G | A | 0.01830471 | ENSEMBL | 55063514 | TRUE |
| rs28362286 | 1 | A | C | -0.2822594 | ENSEMBL | 55063542 | TRUE |
| rs12079951 | 1 | C | A | 0.01610803 | ENSEMBL | 55074653 | TRUE |
| rs571783678 | 1 | T | C | -0.0654458 | ENSEMBL | 55115289 | TRUE |
| rs114022594 | 1 | G | A | -0.0334309 | ENSEMBL | 55123799 | TRUE |
| rs187148846 | 1 | C | T | 0.0626154 | ENSEMBL | 55124560 | TRUE |
| rs61226482 | 1 | A | G | -0.0539748 | ENSEMBL | 55153335 | TRUE |
| rs17111688 | 1 | A | G | 0.02775618 | ENSEMBL | 55161003 | TRUE |
| rs1165221 | 1 | A | G | -0.0161093 | ENSEMBL | 55171947 | TRUE |
| rs72662510 | 1 | T | G | 0.03035359 | ENSEMBL | 55210626 | TRUE |
| rs112334489 | 1 | C | T | -0.0379959 | ENSEMBL | 55214701 | TRUE |
| rs67175330 | 1 | T | C | 0.02220325 | ENSEMBL | 55223088 | TRUE |
| rs575999011 | 1 | T | C | -0.0510932 | ENSEMBL | 55230154 | TRUE |
| rs6684298 | 1 | G | T | -0.0169065 | ENSEMBL | 55251441 | TRUE |
| rs112305482 | 1 | C | T | -0.0159511 | ENSEMBL | 55258644 | TRUE |
| rs2647282 | 1 | C | A | 0.00293306 | ENSEMBL | 55258764 | TRUE |
| rs371443252 | 1 | T | C | -0.2177019 | ENSEMBL | 55260801 | TRUE |
| rs74075412 | 1 | A | G | -0.0315454 | ENSEMBL | 55285240 | TRUE |
| rs116349554 | 1 | T | C | -0.0397932 | ENSEMBL | 55318019 | TRUE |
| rs535066747 | 1 | C | T | -0.2278985 | ENSEMBL | 55337168 | TRUE |
| rs72904635 | 1 | T | C | -0.0151452 | ENSEMBL | 55364140 | TRUE |
| rs2746688 | 1 | G | T | -0.0177833 | ENSEMBL | 55366005 | TRUE |
| rs116182692 | 1 | G | A | -0.1327269 | ENSEMBL | 55499125 | TRUE |
| rs546869935 | 1 | T | C | -0.2576548 | ENSEMBL | 55589223 | TRUE |
| rs148939072 | 1 | C | A | -0.0471834 | ENSEMBL | 55661800 | TRUE |
| rs12091574 | 1 | A | G | -0.0290631 | ENSEMBL | 55713031 | TRUE |
| rs7553801 | 1 | G | A | 0.0160512 | ENSEMBL | 55961727 | TRUE |
| rs111446716 | 1 | T | C | -0.0398354 | ENSEMBL | 62404174 | TRUE |
| rs72913238 | 1 | A | C | -0.0345215 | ENSEMBL | 62489563 | TRUE |
| rs1168018 | 1 | C | A | 0.04061513 | ENSEMBL | 62535449 | TRUE |
| rs995000 | 1 | T | C | -0.0649758 | ENSEMBL | 62641855 | TRUE |
| rs67537755 | 1 | A | G | 0.04299852 | ENSEMBL | 62651657 | TRUE |
| rs7533354 | 1 | C | A | -0.0352361 | ENSEMBL | 62751832 | TRUE |
| rs6661082 | 1 | C | A | -0.0334842 | ENSEMBL | 62786702 | TRUE |
| rs58234250 | 1 | G | T | -0.0253845 | ENSEMBL | 62905739 | TRUE |
| rs114464352 | 1 | C | T | 0.0260578 | ENSEMBL | 92480739 | TRUE |
| rs115885410 | 1 | T | C | 0.02390266 | ENSEMBL | 92741434 | TRUE |
| rs6693893 | 1 | C | T | -0.0272583 | ENSEMBL | 109255141 | TRUE |
| rs592360 | 1 | T | C | 0.01138214 | ENSEMBL | 109259778 | TRUE |
| rs4970834 | 1 | T | C | -0.015673 | ENSEMBL | 109272258 | TRUE |
| rs79868705 | 1 | A | G | -0.0321298 | ENSEMBL | 109272452 | TRUE |
| rs11102967 | 1 | T | C | 0.01703007 | ENSEMBL | 109274623 | TRUE |
| rs12740374 | 1 | T | G | -0.0451519 | ENSEMBL | 109274968 | TRUE |
| rs11577931 | 1 | G | A | -0.0763802 | ENSEMBL | 109278262 | TRUE |
| rs12095374 | 1 | A | G | -0.0215351 | ENSEMBL | 109282257 | TRUE |
| rs138361368 | 1 | A | C | -0.0347729 | ENSEMBL | 109282936 | TRUE |
| rs10910476 | 1 | T | C | 0.01176882 | ENSEMBL | 234599210 | TRUE |
| rs493072 | 2 | T | C | 0.00727737 | ENSEMBL | 16795155 | TRUE |
| rs2114352 | 2 | G | A | 0.02443769 | ENSEMBL | 17641408 | TRUE |
| rs2555103 | 2 | C | T | 0.00679715 | ENSEMBL | 17657334 | TRUE |
| rs13420742 | 2 | G | A | 0.01241063 | ENSEMBL | 18902401 | TRUE |
| rs907866 | 2 | A | G | -0.0161642 | ENSEMBL | 20171619 | TRUE |
| rs79844637 | 2 | T | C | 0.0449213 | ENSEMBL | 20193214 | TRUE |
| rs73916850 | 2 | T | C | -0.0084781 | ENSEMBL | 20532097 | TRUE |
| rs3072 | 2 | C | T | 0.01965659 | ENSEMBL | 20678646 | TRUE |
| rs601325 | 2 | C | T | -0.0140181 | ENSEMBL | 20782631 | TRUE |
| rs6722139 | 2 | A | C | -0.0034528 | ENSEMBL | 20901622 | TRUE |
| rs1318006 | 2 | A | G | -0.0058471 | ENSEMBL | 20991280 | TRUE |
| rs140943736 | 2 | G | T | -0.0187352 | ENSEMBL | 21001236 | TRUE |
| rs12720816 | 2 | C | T | 0.04164684 | ENSEMBL | 21016795 | TRUE |
| rs12713956 | 2 | G | A | -0.0114859 | ENSEMBL | 21018633 | TRUE |
| rs9282603 | 2 | G | A | 0.04818463 | ENSEMBL | 21041014 | TRUE |
| rs1800481 | 2 | A | G | -0.0291267 | ENSEMBL | 21044338 | TRUE |
| rs512535 | 2 | C | T | 0.01364216 | ENSEMBL | 21044910 | TRUE |
| rs79452506 | 2 | A | G | -0.0217739 | ENSEMBL | 21045771 | TRUE |
| rs563290 | 2 | A | G | 0.0316698 | ENSEMBL | 21065354 | TRUE |
| rs571468 | 2 | C | A | 0.02258284 | ENSEMBL | 21071042 | TRUE |
| rs145092252 | 2 | T | C | 0.05100755 | ENSEMBL | 21158898 | TRUE |
| rs76935526 | 2 | T | C | 0.03502278 | ENSEMBL | 21163481 | TRUE |
| rs503662 | 2 | C | T | 0.02894985 | ENSEMBL | 21191270 | TRUE |
| rs553448422 | 2 | C | T | -0.0322262 | ENSEMBL | 21196630 | TRUE |
| rs10198972 | 2 | A | G | -0.0098705 | ENSEMBL | 21230339 | TRUE |
| rs312045 | 2 | T | C | 0.01339107 | ENSEMBL | 21241114 | TRUE |
| rs6733564 | 2 | A | G | 0.01154481 | ENSEMBL | 21256914 | TRUE |
| rs4665662 | 2 | T | G | 0.00539294 | ENSEMBL | 21278472 | TRUE |
| rs10185624 | 2 | G | A | -0.0295061 | ENSEMBL | 21292708 | TRUE |
| rs7605009 | 2 | A | G | -0.0020369 | ENSEMBL | 21317527 | TRUE |
| rs13417640 | 2 | T | C | -0.05248 | ENSEMBL | 21327588 | TRUE |
| rs882967 | 2 | A | C | 0.01370506 | ENSEMBL | 21347847 | TRUE |
| rs635141 | 2 | T | C | 0.01151273 | ENSEMBL | 21358592 | TRUE |
| rs6545834 | 2 | T | C | -0.0171144 | ENSEMBL | 21360230 | TRUE |
| rs186888069 | 2 | A | G | -0.0266598 | ENSEMBL | 21379058 | TRUE |
| rs2164054 | 2 | C | T | 0.00352198 | ENSEMBL | 21389755 | TRUE |
| rs6711455 | 2 | G | A | 0.01407212 | ENSEMBL | 21421132 | TRUE |
| rs2194758 | 2 | A | G | 0.01453364 | ENSEMBL | 21483233 | TRUE |
| rs2946565 | 2 | C | T | 0.02267863 | ENSEMBL | 21944399 | TRUE |
| rs2579950 | 2 | A | G | 0.01801118 | ENSEMBL | 21989613 | TRUE |
| rs6723676 | 2 | A | C | 0.00668003 | ENSEMBL | 22192106 | TRUE |
| rs12478557 | 2 | G | A | 0.01101547 | ENSEMBL | 22256396 | TRUE |
| rs58184964 | 2 | A | G | 0.02472317 | ENSEMBL | 22336004 | TRUE |
| rs1995812 | 2 | A | G | 0.02055297 | ENSEMBL | 22346497 | TRUE |
| rs13007912 | 2 | C | A | 0.01716656 | ENSEMBL | 22564258 | TRUE |
| rs11889451 | 2 | C | T | 0.00216226 | ENSEMBL | 22792413 | TRUE |
| rs2879543 | 2 | C | T | 0.01097027 | ENSEMBL | 23015473 | TRUE |
| rs13015234 | 2 | T | C | 0.02563784 | ENSEMBL | 23389651 | TRUE |
| rs60425099 | 2 | G | A | 0.00983412 | ENSEMBL | 23479515 | TRUE |
| rs4665603 | 2 | G | A | 0.01224676 | ENSEMBL | 23535622 | TRUE |
| rs7603434 | 2 | A | G | 0.01031868 | ENSEMBL | 24509321 | TRUE |
| rs6546148 | 2 | A | G | 0.00567757 | ENSEMBL | 25402136 | TRUE |
| rs7566623 | 2 | G | A | 0.03031663 | ENSEMBL | 26175455 | TRUE |
| rs3828255 | 2 | A | G | 0.05603293 | ENSEMBL | 26190383 | TRUE |
| rs13025681 | 2 | C | T | 0.013899 | ENSEMBL | 26525636 | TRUE |
| rs1141313 | 2 | A | G | 0.02635311 | ENSEMBL | 27238100 | TRUE |
| rs68043976 | 2 | C | T | -0.0327772 | ENSEMBL | 27270768 | TRUE |
| rs780090 | 2 | T | C | 0.11569595 | ENSEMBL | 27495607 | TRUE |
| rs1260326 | 2 | T | C | 0.08899401 | ENSEMBL | 27508073 | TRUE |
| rs4361084 | 2 | C | T | 0.04631577 | ENSEMBL | 27839529 | TRUE |
| rs2063018 | 2 | G | A | 0.03633667 | ENSEMBL | 28095141 | TRUE |
| rs114780578 | 2 | C | A | 0.03952777 | ENSEMBL | 43813316 | TRUE |
| rs4245791 | 2 | C | T | 0.00947885 | ENSEMBL | 43847292 | TRUE |
| rs530169193 | 2 | T | G | 0.14504683 | ENSEMBL | 126383957 | TRUE |
| rs17029617 | 3 | A | G | -0.0159779 | ENSEMBL | 32500061 | TRUE |
| rs145144188 | 4 | G | A | 0.05655128 | ENSEMBL | 68482380 | TRUE |
| rs2878419 | 5 | T | C | 0.00086551 | ENSEMBL | 75344665 | TRUE |
| rs11948514 | 5 | G | A | 0.01470051 | ENSEMBL | 156718845 | TRUE |
| rs10039074 | 5 | T | C | -0.048381 | ENSEMBL | 156949265 | TRUE |
| rs61744066 | 5 | A | G | 0.05885614 | ENSEMBL | 156951655 | TRUE |
| rs12517431 | 5 | T | C | 0.02261031 | ENSEMBL | 156965237 | TRUE |
| rs112812325 | 5 | T | G | 0.02973341 | ENSEMBL | 156988066 | TRUE |
| rs35062987 | 6 | T | C | 0.02033565 | ENSEMBL | 32622958 | TRUE |
| rs3850659 | 6 | T | C | 0.00932644 | ENSEMBL | 160277794 | TRUE |
| rs9457905 | 6 | T | C | 0.020886 | ENSEMBL | 160335898 | TRUE |
| rs146977175 | 6 | A | G | 0.04055706 | ENSEMBL | 160421591 | TRUE |
| rs41272098 | 6 | G | A | 0.03499748 | ENSEMBL | 160587041 | TRUE |
| rs146703276 | 6 | T | C | 0.0541239 | ENSEMBL | 160672592 | TRUE |
| rs4252124 | 6 | T | G | 0.04179355 | ENSEMBL | 160730989 | TRUE |
| rs192891956 | 6 | T | G | 0.03409794 | ENSEMBL | 160870425 | TRUE |
| rs73259906 | 7 | T | C | -0.0328872 | ENSEMBL | 984483 | TRUE |
| rs884978 | 7 | A | G | 0.01778396 | ENSEMBL | 1026423 | TRUE |
| rs77375741 | 7 | A | G | -0.0400355 | ENSEMBL | 1049911 | TRUE |
| rs192361639 | 7 | T | G | 0.00083791 | ENSEMBL | 25951402 | TRUE |
| rs75111196 | 7 | G | A | -0.005902 | ENSEMBL | 137810949 | TRUE |
| rs73729085 | 7 | G | A | -5.83E-05 | ENSEMBL | 137877115 | TRUE |
| rs4841132 | 8 | A | G | 0.01940442 | ENSEMBL | 9326086 | TRUE |
| rs3780181 | 9 | G | A | -0.0229967 | ENSEMBL | 2640759 | TRUE |
| rs4743046 | 9 | T | C | -0.0185639 | ENSEMBL | 96676027 | TRUE |
| rs6477710 | 9 | G | T | -0.0178542 | ENSEMBL | 96823032 | TRUE |
| rs73504341 | 9 | G | A | 0.03733162 | ENSEMBL | 104343252 | TRUE |
| rs193173261 | 9 | G | T | 0.01915792 | ENSEMBL | 104719670 | TRUE |
| rs62565985 | 9 | G | A | -0.0265658 | ENSEMBL | 104734525 | TRUE |
| rs2417562 | 9 | G | A | -0.029449 | ENSEMBL | 104747036 | TRUE |
| rs77877520 | 9 | G | A | 0.03425192 | ENSEMBL | 104781232 | TRUE |
| rs4149307 | 9 | C | T | -0.0200424 | ENSEMBL | 104827463 | TRUE |
| rs2253175 | 9 | C | T | -0.0160114 | ENSEMBL | 104858018 | TRUE |
| rs418057 | 9 | C | T | -0.0266009 | ENSEMBL | 105288663 | TRUE |
| rs16924584 | 9 | G | T | 0.02517944 | ENSEMBL | 105416834 | TRUE |
| rs10759180 | 9 | G | A | -0.0284943 | ENSEMBL | 106400535 | TRUE |
| rs2098272 | 9 | C | T | -0.0166296 | ENSEMBL | 120134844 | TRUE |
| rs11239541 | 10 | T | C | 0.010752 | ENSEMBL | 45506418 | TRUE |
| rs77150189 | 10 | C | A | -0.0190885 | ENSEMBL | 112153205 | TRUE |
| rs5095 | 11 | G | A | -0.0760792 | ENSEMBL | 116822447 | TRUE |
| rs12721092 | 11 | C | T | -0.0526906 | ENSEMBL | 116829478 | TRUE |
| rs17120099 | 11 | C | A | -0.1076043 | ENSEMBL | 116864744 | TRUE |
| rs17030221 | 12 | T | C | -0.0133116 | ENSEMBL | 100499570 | TRUE |
| rs62001736 | 15 | A | G | 0.01570147 | ENSEMBL | 58381852 | TRUE |
| rs261291 | 15 | C | T | 0.00690907 | ENSEMBL | 58387979 | TRUE |
| rs174418 | 15 | C | T | -0.0077907 | ENSEMBL | 58395404 | TRUE |
| rs723967 | 15 | C | T | -0.0111818 | ENSEMBL | 58404101 | TRUE |
| rs1800588 | 15 | C | T | -0.0179079 | ENSEMBL | 58431476 | TRUE |
| rs148562851 | 16 | G | A | -0.0536889 | ENSEMBL | 56960247 | TRUE |
| rs4783961 | 16 | A | G | -0.0403662 | ENSEMBL | 56960982 | TRUE |
| rs17231520 | 16 | A | G | -0.0696791 | ENSEMBL | 56961915 | TRUE |
| rs7499892 | 16 | T | C | 0.02830566 | ENSEMBL | 56972678 | TRUE |
| rs289719 | 16 | T | C | -0.0060386 | ENSEMBL | 56976029 | TRUE |
| rs9926649 | 16 | T | G | -0.0053181 | ENSEMBL | 66716137 | TRUE |
| rs111810144 | 16 | T | C | -0.0087761 | ENSEMBL | 67182207 | TRUE |
| rs9933206 | 16 | T | C | -0.0073336 | ENSEMBL | 67505119 | TRUE |
| rs58624196 | 16 | A | G | -0.0042023 | ENSEMBL | 67549826 | TRUE |
| rs571677130 | 16 | T | C | -0.0054152 | ENSEMBL | 67601810 | TRUE |
| rs9934405 | 16 | C | A | -0.0007396 | ENSEMBL | 67956073 | TRUE |
| rs187285131 | 16 | A | G | 0.00529687 | ENSEMBL | 68289493 | TRUE |
| rs116747877 | 16 | A | G | 0.00973706 | ENSEMBL | 68319916 | TRUE |
| rs12934168 | 16 | T | C | -0.0104739 | ENSEMBL | 71402875 | TRUE |
| rs8060083 | 16 | T | C | 0.00475527 | ENSEMBL | 71416256 | TRUE |
| rs146413278 | 16 | T | C | 0.01060735 | ENSEMBL | 71742865 | TRUE |
| rs145946926 | 16 | T | C | 0.0886931 | ENSEMBL | 71906842 | TRUE |
| rs142376190 | 16 | T | C | 0.05357994 | ENSEMBL | 71981199 | TRUE |
| rs114374667 | 16 | A | G | 0.01773792 | ENSEMBL | 71989659 | TRUE |
| rs35697801 | 16 | C | T | -0.0111485 | ENSEMBL | 71994321 | TRUE |
| rs9938539 | 16 | C | T | -0.0086751 | ENSEMBL | 71996681 | TRUE |
| rs5471 | 16 | C | A | 0.03823208 | ENSEMBL | 72054562 | TRUE |
| rs16973703 | 16 | T | C | 0.00230461 | ENSEMBL | 72086784 | TRUE |
| rs146826822 | 16 | T | C | 0.09682756 | ENSEMBL | 72175911 | TRUE |
| rs7196334 | 16 | G | T | 0.01023922 | ENSEMBL | 72676786 | TRUE |
| rs2526068 | 16 | G | A | -0.0118873 | ENSEMBL | 74001887 | TRUE |
| rs11868705 | 17 | A | G | -0.0384398 | ENSEMBL | 4789440 | TRUE |
| rs3744841 | 18 | G | A | 0.00589971 | ENSEMBL | 49591004 | TRUE |
| rs138847616 | 19 | C | T | -0.1177736 | ENSEMBL | 10148588 | TRUE |
| rs139501553 | 19 | C | T | -0.1114675 | ENSEMBL | 10278711 | TRUE |
| rs114810281 | 19 | C | T | -0.0496394 | ENSEMBL | 10305136 | TRUE |
| rs533063641 | 19 | T | C | -0.1006203 | ENSEMBL | 10347633 | TRUE |
| rs12720261 | 19 | T | C | -0.1162504 | ENSEMBL | 10367145 | TRUE |
| rs111317755 | 19 | T | C | -0.0123964 | ENSEMBL | 10496495 | TRUE |
| rs114690859 | 19 | T | C | -0.0377719 | ENSEMBL | 10503112 | TRUE |
| rs74695382 | 19 | C | T | -0.0793237 | ENSEMBL | 10548704 | TRUE |
| rs535500504 | 19 | T | C | -0.1600584 | ENSEMBL | 10740461 | TRUE |
| rs375200581 | 19 | G | A | -0.0400518 | ENSEMBL | 10795047 | TRUE |
| rs190035797 | 19 | A | G | -0.1373038 | ENSEMBL | 10839261 | TRUE |
| rs577964300 | 19 | T | G | 0.04466234 | ENSEMBL | 10888539 | TRUE |
| rs142783157 | 19 | A | G | -0.0256584 | ENSEMBL | 10917338 | TRUE |
| rs77611468 | 19 | G | A | 0.01451483 | ENSEMBL | 10989785 | TRUE |
| rs138911317 | 19 | A | G | -0.0267393 | ENSEMBL | 10992834 | TRUE |
| rs3786721 | 19 | T | C | 0.00347905 | ENSEMBL | 11035823 | TRUE |
| rs189594619 | 19 | G | A | -0.1985224 | ENSEMBL | 11044139 | TRUE |
| rs559108663 | 19 | C | A | 0.04139152 | ENSEMBL | 11050471 | TRUE |
| rs55677033 | 19 | T | C | 0.01418516 | ENSEMBL | 11055617 | TRUE |
| rs113365523 | 19 | T | C | -0.0228224 | ENSEMBL | 11066195 | TRUE |
| rs144844033 | 19 | T | C | -0.0206261 | ENSEMBL | 11067195 | TRUE |
| rs114601270 | 19 | A | G | 0.02567795 | ENSEMBL | 11067303 | TRUE |
| rs9305019 | 19 | T | C | -0.0177849 | ENSEMBL | 11075736 | TRUE |
| rs9305020 | 19 | T | C | 0.01167622 | ENSEMBL | 11076035 | TRUE |
| rs139027728 | 19 | C | A | 0.03832985 | ENSEMBL | 11080920 | TRUE |
| rs138294113 | 19 | T | C | -0.0386953 | ENSEMBL | 11081053 | TRUE |
| rs140346950 | 19 | A | G | -0.0461597 | ENSEMBL | 11085265 | TRUE |
| rs114197570 | 19 | T | C | -0.1565686 | ENSEMBL | 11085594 | TRUE |
| rs560828472 | 19 | C | A | -0.2004772 | ENSEMBL | 11093461 | TRUE |
| rs3745678 | 19 | T | C | -0.0546893 | ENSEMBL | 11100403 | TRUE |
| rs59078413 | 19 | C | T | 0.03029593 | ENSEMBL | 11102036 | TRUE |
| rs74257940 | 19 | G | T | 0.04249804 | ENSEMBL | 11103799 | TRUE |
| rs11669576 | 19 | A | G | 0.01656965 | ENSEMBL | 11111624 | TRUE |
| rs17248833 | 19 | G | A | -0.0179696 | ENSEMBL | 11113916 | TRUE |
| rs190729088 | 19 | A | G | -0.1529829 | ENSEMBL | 11114343 | TRUE |
| rs1799898 | 19 | T | C | -0.0293031 | ENSEMBL | 11116878 | TRUE |
| rs146957693 | 19 | T | C | -0.0081547 | ENSEMBL | 11124635 | TRUE |
| rs11670740 | 19 | G | A | 0.01042869 | ENSEMBL | 11143956 | TRUE |
| rs149442147 | 19 | G | A | -0.0439583 | ENSEMBL | 11146568 | TRUE |
| rs143813975 | 19 | A | G | -0.0296625 | ENSEMBL | 11164046 | TRUE |
| rs4804573 | 19 | G | A | 0.02161085 | ENSEMBL | 11166556 | TRUE |
| rs4804574 | 19 | G | A | 0.01312349 | ENSEMBL | 11206806 | TRUE |
| rs17699030 | 19 | G | A | -0.034498 | ENSEMBL | 11220266 | TRUE |
| rs10406522 | 19 | T | C | -0.0097274 | ENSEMBL | 11230959 | TRUE |
| rs737338 | 19 | T | C | -0.0285179 | ENSEMBL | 11236981 | TRUE |
| rs141409550 | 19 | A | G | -0.0287655 | ENSEMBL | 11246556 | TRUE |
| rs76204556 | 19 | T | C | -0.0645317 | ENSEMBL | 11282659 | TRUE |
| rs79675274 | 19 | T | C | -0.0017222 | ENSEMBL | 11294058 | TRUE |
| rs140028947 | 19 | G | A | -0.0774423 | ENSEMBL | 11321261 | TRUE |
| rs552638138 | 19 | A | G | -0.0354243 | ENSEMBL | 11589134 | TRUE |
| rs142527337 | 19 | A | G | 0.03725065 | ENSEMBL | 44799635 | TRUE |
| rs28399641 | 19 | T | C | -0.0345782 | ENSEMBL | 44821701 | TRUE |
| rs78330310 | 19 | A | G | -0.0633834 | ENSEMBL | 44858385 | TRUE |
| rs73556103 | 19 | T | C | -0.0318124 | ENSEMBL | 44870078 | TRUE |
| rs186930841 | 19 | T | C | -0.2248337 | ENSEMBL | 44878201 | TRUE |
| rs34224078 | 19 | G | A | -0.0691213 | ENSEMBL | 44879858 | TRUE |
| rs149132410 | 19 | A | G | -0.1594222 | ENSEMBL | 44880231 | TRUE |
| rs78754926 | 19 | A | G | -0.0422988 | ENSEMBL | 44881845 | TRUE |
| rs166907 | 19 | G | A | 0.05675135 | ENSEMBL | 44883598 | TRUE |
| rs2075649 | 19 | G | A | 0.00731243 | ENSEMBL | 44892073 | TRUE |
| rs28480204 | 19 | T | G | 0.05646848 | ENSEMBL | 44894086 | TRUE |
| rs157589 | 19 | A | G | 0.08374998 | ENSEMBL | 44895062 | TRUE |
| rs190073745 | 19 | G | A | 0.02399176 | ENSEMBL | 44901792 | TRUE |
| rs769446 | 19 | C | T | 0.06170932 | ENSEMBL | 44905371 | TRUE |
| rs405509 | 19 | T | G | 0.02432799 | ENSEMBL | 44905579 | TRUE |
| rs429358 | 19 | C | T | 0.06613555 | ENSEMBL | 44908684 | TRUE |
| rs5114 | 19 | T | C | -0.2024673 | ENSEMBL | 44915189 | TRUE |
| rs540950376 | 19 | G | A | -0.232155 | ENSEMBL | 44918507 | TRUE |
| rs12721054 | 19 | G | A | -0.2062004 | ENSEMBL | 44919330 | TRUE |
| rs145713584 | 19 | A | G | 0.07376225 | ENSEMBL | 44922170 | TRUE |
| rs114498232 | 19 | T | C | 0.0521646 | ENSEMBL | 44929745 | TRUE |
| rs370625306 | 19 | T | C | -0.226454 | ENSEMBL | 44931624 | TRUE |
| rs140846591 | 19 | C | T | -0.1488375 | ENSEMBL | 44939877 | TRUE |
| rs34648425 | 19 | G | A | 0.00341496 | ENSEMBL | 45004697 | TRUE |
| rs78049117 | 19 | A | G | -0.0238413 | ENSEMBL | 45016628 | TRUE |
| rs111544400 | 19 | C | T | -0.0511548 | ENSEMBL | 45031122 | TRUE |
| rs143394534 | 19 | T | C | -0.065435 | ENSEMBL | 45206085 | TRUE |
| rs56845483 | 19 | C | T | -0.0239629 | ENSEMBL | 45210040 | TRUE |
| rs8116470 | 20 | A | G | -0.0261717 | ENSEMBL | 35579460 | TRUE |
| rs2836926 | 21 | A | G | 0.012168 | ENSEMBL | 39164864 | TRUE |
| rs77974343 | 21 | T | C | -0.2134086 | ENSEMBL | 45496290 | TRUE |

**Supplemental Table S4. Potentially Causal Variants for Triglycerides Identified by the Global Lipids Genetic Consortium.**(7)

| **Chromosome** | **Position (GRCh37)** | **A1** | **A2** | **Notes** |
| --- | --- | --- | --- | --- |
| 1 | 27021913 | G | C |  |
| 1 | 27284913 | C | T |  |
| 1 | 54890956 | T | G |  |
| 1 | 61680217 | A | G | Not available in AoU |
| 1 | 61994357 | C | T | Not available in AoU |
| 1 | 62920008 | A | G |  |
| 1 | 62957030 | G | A |  |
| 1 | 63070537 | AGTTAATGTG | A |  |
| 1 | 63107526 | C | T | Also in the PRS |
| 1 | 64576995 | C | T |  |
| 1 | 184865132 | T | A |  |
| 1 | 220970028 | A | G |  |
| 1 | 230294715 | C | A |  |
| 2 | 21231524 | G | A |  |
| 2 | 21383353 | C | T |  |
| 2 | 21385778 | C | G |  |
| 2 | 27393030 | A | G |  |
| 2 | 27399294 | C | T |  |
| 2 | 27730940 | T | C | Also in the PRS |
| 2 | 219699999 | G | A |  |
| 2 | 227107501 | C | T |  |
| 2 | 234074745 | C | T |  |
| 3 | 24520283 | A | G |  |
| 3 | 87037543 | A | G |  |
| 3 | 150066540 | T | A |  |
| 3 | 172294500 | G | A |  |
| 4 | 951947 | T | C |  |
| 4 | 3287052 | C | A |  |
| 4 | 3387148 | C | T |  |
| 4 | 3443931 | A | G |  |
| 4 | 4990298 | A | G |  |
| 4 | 26047616 | A | G |  |
| 4 | 87772240 | T | C |  |
| 4 | 103188709 | C | T |  |
| 4 | 110578226 | T | A |  |
| 4 | 110638824 | C | T |  |
| 4 | 155489608 | C | T |  |
| 5 | 67714246 | A | G |  |
| 5 | 131008194 | T | C |  |
| 5 | 132444128 | G | A |  |
| 5 | 156391628 | T | C |  |
| 6 | 31270118 | C | A |  |
| 6 | 31326289 | G | A |  |
| 6 | 33761462 | T | C |  |
| 6 | 40998167 | T | C |  |
| 6 | 43757896 | C | A |  |
| 6 | 127440047 | T | C |  |
| 6 | 160964135 | T | C |  |
| 6 | 161008646 | G | A |  |
| 6 | 161092438 | C | T |  |
| 7 | 25991826 | T | C |  |
| 7 | 73020337 | C | G |  |
| 8 | 19742204 | T | A |  |
| 8 | 19820916 | C | T |  |
| 8 | 19830921 | C | T |  |
| 8 | 20118438 | C | T | Not available in AoU |
| 8 | 126137401 | G | A |  |
| 8 | 126500031 | C | G |  |
| 9 | 16901067 | C | A |  |
| 9 | 102162570 | C | T |  |
| 9 | 107665978 | C | G |  |
| 9 | 117083803 | C | A |  |
| 10 | 52573772 | C | T |  |
| 10 | 65191645 | G | T |  |
| 10 | 94530832 | A | C |  |
| 10 | 94843535 | T | C |  |
| 10 | 134459388 | A | G |  |
| 11 | 14451992 | T | C |  |
| 11 | 14865399 | T | C |  |
| 11 | 61588305 | A | G |  |
| 11 | 64004723 | G | C |  |
| 11 | 65473798 | C | A |  |
| 11 | 114739047 | C | T |  |
| 11 | 114913567 | T | G |  |
| 11 | 116623213 | TA | T |  |
| 11 | 116648917 | G | C |  |
| 11 | 116662407 | G | C | Also among variants in the 5 canonical TG genes |
| 11 | 117000856 | T | TG |  |
| 11 | 117018764 | T | G |  |
| 11 | 117053959 | G | A |  |
| 11 | 117491209 | C | T |  |
| 11 | 120068136 | C | T |  |
| 12 | 4384844 | T | G |  |
| 12 | 21331549 | T | C |  |
| 12 | 48143315 | A | G |  |
| 12 | 49399132 | G | C |  |
| 12 | 109661672 | A | G |  |
| 12 | 125312425 | G | C |  |
| 13 | 45970147 | A | G |  |
| 13 | 114551993 | T | C |  |
| 14 | 38848824 | T | A |  |
| 14 | 50655357 | G | C |  |
| 14 | 100765823 | T | C |  |
| 15 | 40397191 | C | G |  |
| 15 | 41057507 | C | T |  |
| 15 | 42371452 | G | A |  |
| 15 | 44027885 | T | C |  |
| 15 | 44581461 | G | A |  |
| 15 | 58723426 | A | G |  |
| 15 | 58855748 | C | T |  |
| 15 | 90214777 | G | A |  |
| 15 | 102068658 | G | A |  |
| 16 | 15150505 | C | A |  |
| 16 | 49886366 | T | C |  |
| 16 | 72108093 | G | A |  |
| 16 | 81534790 | T | C |  |
| 16 | 85150163 | A | G |  |
| 17 | 1618363 | T | C |  |
| 17 | 4692640 | G | T |  |
| 17 | 7106378 | G | A |  |
| 17 | 26694861 | G | A |  |
| 17 | 27889643 | C | T |  |
| 17 | 41874745 | C | A |  |
| 17 | 41926126 | C | T |  |
| 17 | 42155742 | C | T |  |
| 17 | 46197755 | A | G |  |
| 17 | 64210580 | A | C |  |
| 17 | 67081278 | A | G |  |
| 17 | 74268619 | C | T |  |
| 18 | 2846812 | A | T |  |
| 18 | 60845884 | T | C |  |
| 19 | 8336373 | AGGAAGGGAAGG | A |  |
| 19 | 8429323 | G | A |  |
| 19 | 8596372 | ACAG | A |  |
| 19 | 9838421 | G | A |  |
| 19 | 11350874 | C | T |  |
| 19 | 19130750 | G | A |  |
| 19 | 19379549 | C | T |  |
| 19 | 41754430 | G | A |  |
| 19 | 44356388 | T | A | Not available in AoU |
| 19 | 45386229 | A | G |  |
| 19 | 45390333 | A | G |  |
| 19 | 45416178 | G | T |  |
| 19 | 45422587 | A | G | Also in the PRS |
| 19 | 49259529 | A | G |  |
| 20 | 39179822 | G | C |  |
| 20 | 44551855 | T | C |  |
| 20 | 62711459 | C | T |  |
| 21 | 42622479 | A | G |  |
| 21 | 46875775 | G | A |  |
| 21 | 46875817 | G | A | Not available in AoU |
| 21 | 46916204 | C | T | Also in the PRS |
| 22 | 17625915 | G | A |  |
| 22 | 36042986 | C | T |  |
| 22 | 46687681 | G | C |  |

AoU = All of Us Research Program; TG = triglyceride; PRS = Polygenic Risk Score

**Supplemental Table S5. Annotation of Likely Functional Variants within 5 Canonical Triglyceride Gene Regions.**

| **Gene** | **Chr** | **Position** | **rsID** | **Minor** | **Major** | **HTG group carriage^†^** | **Previous citation** | **MAF.1000G** | **MAF** | **NCHROBS** | **CADD.phred** | **Mutation Taster** | **SIFTcat** | **PolyPhenCat** | **ClinVar_Class** |
| --- | --- | --- | --- | --- | --- | --- | --- | --- | --- | --- | --- | --- | --- | --- | --- |
| *LPL* | 8 | 19939489 | rs756418111 | G | A |  |  | - | 3.25E-05 | 30744 | 20.5 | 0.999 | tolerated | benign | not in ClinVar |
| *LPL* | 8 | 19939492 | rs1259299905 | G | C |  |  | - | 6.51E-05 | 30746 | 22.2 | 0.976 | tolerated | probably_damaging | Likely benign |
| *LPL* | 8 | 19939499 | rs572477224 | T | C |  |  | 0 | 6.51E-05 | 30746 | 17.4 | 1 | tolerated | benign | Likely benign |
| *LPL* | 8 | 19939511 | rs761167661 | C | G |  |  | - | 3.25E-05 | 30746 | 18.33 | 0.976 | tolerated | benign | not in ClinVar |
| *LPL* | 8 | 19948188 | rs2069900175 | C | G |  |  |  | 3.25E-05 | 30746 | 23.3 | 0.776 | deleterious | probably_damaging | not in ClinVar |
| *LPL* | 8 | 19948200 | rs538543355 | G | A |  |  | 8.00E-04 | 1.95E-04 | 30744 | 22.6 | 1 | tolerated | possibly_damaging | not in ClinVar |
| *LPL* | 8 | 19948219 | rs1423027681 | A | T |  |  | - | 3.25E-05 | 30740 | 24 | 0.999 | deleterious | probably_damaging | Uncertain significance |
| *LPL* | 8 | 19948233 | rs1015492279 | T | G |  |  | - | 6.51E-05 | 30746 | 16.92 | 0.987 | tolerated | benign | not in ClinVar |
| *LPL* | 8 | 19948240 | rs148201569 | G | C | 1 |  | 4.50E-03 | 2.34E-03 | 30746 | 14.15 | 0.995 | tolerated | benign | not in ClinVar |
| *LPL* | 8 | 19948240 | rs148201569 | T | C |  |  | - | 6.51E-05 | 30746 | 14.03 | 0.994 | tolerated | benign | not in ClinVar |
| *LPL* | 8 | 19948281 | rs114101772 | A | G |  | S4, SD | 0 | 3.25E-05 | 30746 | 18.82 | 0.763 | deleterious | possibly_damaging | Uncertain significance |
| *LPL* | 8 | 19948339 | rs372267210 | T | C |  |  | - | 6.51E-05 | 30746 | 24.3 | 1 | deleterious | probably_damaging | Uncertain significance |
| *LPL* | 8 | 19951776 | rs2069936138 | C | G |  |  |  | 3.25E-05 | 30744 | 27.6 | 1 | deleterious | probably_damaging | Uncertain significance |
| *LPL* | 8 | 19951778 | rs1240452664 | T | A |  |  | - | 3.25E-05 | 30736 | 23.1 | 1 | tolerated | benign | not in ClinVar |
| *LPL* | 8 | 19951805 | rs373088068 | C | G |  | S2, SD | - | 6.51E-05 | 30744 | 27.9 | 1 | deleterious | probably_damaging | Conflicting classifications of pathogenicity |
| *LPL* | 8 | 19951823 | NA | G | A |  |  |  | 3.25E-05 | 30744 | 22.4 | 1 | deleterious | possibly_damaging | not in ClinVar |
| *LPL* | 8 | 19951895 | NA | A | G |  |  |  | 3.25E-05 | 30746 | 13.15 | 1 | tolerated | benign | not in ClinVar |
| *LPL* | 8 | 19951905 | rs140903633 | C | A |  |  | - | 6.51E-05 | 30740 | 25.5 | 0.788 | deleterious | probably_damaging | Uncertain significance |
| *LPL* | 8 | 19951928 | rs376875031 | T | C |  |  | - | 1.30E-04 | 30746 | 20.4 | 1 | deleterious | possibly_damaging | not in ClinVar |
| *LPL* | 8 | 19951929 | rs527267420 | A | G |  |  | - | 6.51E-05 | 30746 | 13.13 | 1 | tolerated | benign | Uncertain significance |
| *LPL* | 8 | 19953337 | rs748459586 | C | G |  |  | - | 3.25E-05 | 30744 | 11.84 | 0.909 | tolerated | benign | not in ClinVar |
| *LPL* | 8 | 19953389 | rs753632194 | A | G |  |  | - | 9.76E-05 | 30746 | 13.22 | 0.998 | tolerated | benign | not in ClinVar |
| *LPL* | 8 | 19953406 | NA | C | G |  |  |  | 3.25E-05 | 30746 | 23.9 | 0.998 | tolerated | possibly_damaging | not in ClinVar |
| *LPL* | 8 | 19954125 | rs781614031 | A | G | 1 |  | - | 3.25E-05 | 30746 | 27 | 1 | deleterious | probably_damaging | Pathogenic/Likely pathogenic |
| *LPL* | 8 | 19954150 | rs544932321 | T | A |  |  | 3.00E-03 | 4.23E-04 | 30744 | 22.4 | 0.973 | tolerated | benign | Uncertain significance |
| *LPL* | 8 | 19954185 | rs118204056 | A | G |  |  | - | 6.51E-05 | 30744 | 26 | 1 | deleterious | probably_damaging | Pathogenic |
| *LPL* | 8 | 19954222 | rs118204057 | A | G |  | S2, S3, Mild, SD | 0 | 1.30E-04 | 30744 | 25.3 | 1 | tolerated | probably_damaging | Conflicting classifications of pathogenicity |
| *LPL* | 8 | 19954240 | rs118204061 | C | T |  | S3, SD | - | 3.25E-05 | 30728 | 28.4 | 1 | deleterious | probably_damaging | Conflicting classifications of pathogenicity |
| *LPL* | 8 | 19954257 | rs1389582732 | C | G |  |  | - | 3.25E-05 | 30746 | 26 | 1 | deleterious | possibly_damaging | not in ClinVar |
| *LPL* | 8 | 19954258 | rs528243561 | C | T |  | S2 | 0 | 3.25E-05 | 30738 | 27.7 | 1 | deleterious | possibly_damaging | Uncertain significance |
| *LPL* | 8 | 19954279 | rs118204060 | T | C |  | S2, SD | - | 3.25E-05 | 30744 | 27 | 1 | deleterious | probably_damaging | Pathogenic/Likely pathogenic |
| *LPL* | 8 | 19954317 | rs905583712 | A | G |  |  | - | 3.25E-05 | 30746 | 23.7 | 0.957 | tolerated | benign | not in ClinVar |
| *LPL* | 8 | 19954324 | NA | G | T | 1 |  |  | 3.25E-05 | 30742 | 25.1 | 0.77 | deleterious | benign | not in ClinVar |
| *LPL* | 8 | 19955852 | NA | G | C |  |  |  | 3.25E-05 | 30746 | 11.3 | 1 | tolerated | benign | not in ClinVar |
| *LPL* | 8 | 19955939 | rs2069980923 | G | A |  |  |  | 6.51E-05 | 30740 | 16.93 | 0.999 | tolerated | benign | not in ClinVar |
| *LPL* | 8 | 19955976 | rs769886647 | A | G |  |  | - | 3.25E-05 | 30744 | 24.2 | 1 | deleterious | benign | Uncertain significance |
| *LPL* | 8 | 19956005 | rs1489499895 | A | G |  |  | - | 3.25E-05 | 30742 | 26.1 | 1 | tolerated | possibly_damaging | not in ClinVar |
| *LPL* | 8 | 19956018 | rs268 | G | A | 1 | SD | 8.00E-04 | 2.86E-03 | 30746 | 16.12 | 0.989 | tolerated | benign | Conflicting classifications of pathogenicity |
| *LPL* | 8 | 19956029 | NA | A | G |  |  |  | 3.25E-05 | 30746 | 11.81 | 0.999 | tolerated | benign | not in ClinVar |
| *LPL* | 8 | 19956039 | rs1353632906 | C | G |  |  | - | 6.51E-05 | 30744 | 23.3 | 0.753 | deleterious | benign | not in ClinVar |
| *LPL* | 8 | 19956053 | rs2069982720 | G | C | 1 |  |  | 6.51E-05 | 30746 | 16.86 | 0.999 | deleterious | benign | not in ClinVar |
| *LPL* | 8 | 19956063 | rs144466625 | A | G | 1 |  | 8.00E-04 | 9.11E-04 | 30746 | 26.5 | 1 | deleterious | benign | Conflicting classifications of pathogenicity |
| *LPL* | 8 | 19959349 | rs298 | A | G |  |  | - | 3.25E-05 | 30746 | 14.3 | 1 | tolerated | benign | Uncertain significance |
| *LPL* | 8 | 19959359 | NA | A | G |  |  |  | 3.25E-05 | 30744 | 17.26 | 0.646 | tolerated | benign | not in ClinVar |
| *LPL* | 8 | 19959364 | rs780133524 | G | A |  |  | - | 3.25E-05 | 30746 | 22 | 0.946 | tolerated | benign | Uncertain significance |
| *LPL* | 8 | 19960930 | rs141502542 | T | C |  |  | - | 2.60E-04 | 30742 | 24 | 1 | deleterious | benign | not in ClinVar |
| *LPL* | 8 | 19960957 | rs139282874 | C | T |  |  | - | 2.28E-04 | 30744 | 25.9 | 1 | deleterious | possibly_damaging | Uncertain significance |
| *LPL* | 8 | 19960962 | rs150009614 | A | G |  |  | - | 3.25E-05 | 30744 | 29.6 | 0.999 | deleterious | possibly_damaging | Uncertain significance |
| *LPL* | 8 | 19960962 | rs150009614 | C | G |  |  | - | 9.76E-05 | 30744 | 27.6 | 0.996 | deleterious | probably_damaging | Uncertain significance |
| *LPL* | 8 | 19960966 | rs1392526145 | C | T |  |  | - | 9.76E-05 | 30742 | 28.2 | 1 | deleterious | probably_damaging | not in ClinVar |
| *LPL* | 8 | 19960995 | rs541991367 | C | G |  |  | - | 6.51E-05 | 30744 | 19.09 | 0.597 | deleterious | probably_damaging | Uncertain significance |
| *LPL* | 8 | 19961025 | rs1281799804 | G | A |  |  | - | 3.25E-05 | 30744 | 15.79 | 0.502 | tolerated | benign | not in ClinVar |
| *LPL* | 8 | 19961051 | rs767740111 | T | G |  |  | - | 3.25E-05 | 30742 | 22.3 | 0.626 | deleterious | possibly_damaging | Uncertain significance |
| *LPL* | 8 | 19962117 | rs116403115 | G | T |  | S4, SD | 0 | 3.25E-05 | 30746 | 27.4 | 1 | deleterious | possibly_damaging | Conflicting classifications of pathogenicity |
| *LPL* | 8 | 19962134 | rs149089920 | A | G |  |  | 8.00E-04 | 5.53E-04 | 30746 | 24.1 | 0.958 | deleterious | benign | Uncertain significance |
| *LPL* | 8 | 19962163 | rs745596838 | C | G |  |  | - | 3.25E-05 | 30744 | 17.9 | 0.714 | tolerated | possibly_damaging | Uncertain significance |
| *LPL* | 8 | 19962173 | rs1448783719 | A | G |  |  | - | 6.51E-05 | 30744 | 17.17 | 0.927 | tolerated | benign | not in ClinVar |
| *LPL* | 8 | 19962177 | rs776532284 | C | T |  |  | - | 3.25E-05 | 30742 | 29.2 | 1 | deleterious | probably_damaging | Uncertain significance |
| *LPL* | 8 | 19962206 | rs1364598167 | C | A |  |  | - | 1.63E-04 | 30744 | 19.24 | 0.701 | deleterious | benign | Uncertain significance |
| *GPIHBP1* | 8 | 143213284 | NA | G | C |  |  |  | 3.25E-05 | 30744 | 20.5 | 1 | deleterious | possibly_damaging | not in ClinVar |
| *GPIHBP1* | 8 | 143213298 | rs370545732 | T | C |  |  | - | 3.25E-05 | 30746 | 17.27 | 1 | deleterious | probably_damaging | Uncertain significance |
| *GPIHBP1* | 8 | 143213935 | rs587777636 | C | G |  |  | - | 6.51E-05 | 30740 | 19.35 | 1 | deleterious | possibly_damaging | Uncertain significance |
| *GPIHBP1* | 8 | 143215019 | NA | C | T |  |  |  | 3.25E-05 | 30744 | 20.1 | 1 | deleterious | probably_damaging | Uncertain significance |
| *GPIHBP1* | 8 | 143215054 | rs375004793 | A | G |  |  | - | 1.95E-04 | 30746 | 20.9 | 1 | deleterious | possibly_damaging | not in ClinVar |
| *GPIHBP1* | 8 | 143215098 | rs776388699 | A | C |  |  | - | 3.25E-05 | 30746 | 35 | 1 | NA | NA | Pathogenic |
| *GPIHBP1* | 8 | 143215114 | rs1816283022 | T | C | 1 |  |  | 6.51E-05 | 30746 | 11.85 | 1 | deleterious | benign | not in ClinVar |
| *GPIHBP1* | 8 | 143215322 | rs371650837 | T | C |  |  | - | 9.76E-05 | 30746 | 17.06 | 0.818 | deleterious | probably_damaging | not in ClinVar |
| *GPIHBP1* | 8 | 143215324 | rs1216079888 | A | G |  |  | - | 3.25E-05 | 30746 | 12.99 | 1 | tolerated | probably_damaging | not in ClinVar |
| *GPIHBP1* | 8 | 143215342 | rs369222108 | G | A |  |  | - | 6.51E-05 | 30744 | 21.2 | 1 | deleterious | possibly_damaging | not in ClinVar |
| *GPIHBP1* | 8 | 143215358 | rs1313868716 | G | A |  |  | - | 3.25E-05 | 30742 | 17.04 | 0.995 | deleterious | benign | not in ClinVar |
| *GPIHBP1* | 8 | 143215373 | rs1816289317 | G | A |  |  |  | 3.25E-05 | 30736 | 21.2 | 0.939 | deleterious | possibly_damaging | not in ClinVar |
| *GPIHBP1* | 8 | 143215394 | rs78367243 | T | C | 1 | SD | 0.177 | 0.1594* | 30746 | 21.2 | 1 | deleterious | probably_damaging | Benign |
| *GPIHBP1* | 8 | 143215399 | NA | T | G |  |  |  | 3.25E-05 | 30744 | 13.39 | 1 | tolerated | possibly_damaging | not in ClinVar |
| *GPIHBP1* | 8 | 143215432 | rs374384937 | A | C |  |  | - | 3.25E-05 | 30746 | 14.72 | 1 | deleterious | possibly_damaging | not in ClinVar |
| *GPIHBP1* | 8 | 143215486 | rs145844329 | C | G | 1 | S3, SD | 7.60E-03 | 5.85E-03 | 30746 | 16.31 | 1 | deleterious | probably_damaging | Conflicting classifications of pathogenicity |
| *GPIHBP1* | 8 | 143215514 | rs563623646 | T | C |  |  | 8.00E-04 | 3.25E-05 | 30746 | 11.26 | 1 | deleterious | benign | Uncertain significance |
| *APOA5* | 11 | 116790163 | rs1940968954 | A | G |  |  |  | 3.25E-05 | 30746 | 19.83 | 0.999 | deleterious | benign | Uncertain significance |
| *APOA5* | 11 | 116790267 | rs201201147 | A | T | 1 | S4, SD | 0 | 9.76E-05 | 30742 | 24.5 | 0.749 | deleterious | possibly_damaging | Benign/Likely benign |
| *APOA5* | 11 | 116790273 | rs781438417 | A | G |  |  | - | 3.25E-05 | 30742 | 25.4 | 0.998 | deleterious | probably_damaging | not in ClinVar |
| *APOA5* | 11 | 116790285 | rs143292359 | A | G |  | S2, S3, Mild, SD | 8.00E-04 | 3.25E-05 | 30746 | 24.7 | 0.729 | deleterious | possibly_damaging | Benign/Likely benign |
| *APOA5* | 11 | 116790313 | rs1017843978 | T | C |  |  | - | 3.25E-05 | 30746 | 24.1 | 0.989 | tolerated | possibly_damaging | Uncertain significance |
| *APOA5* | 11 | 116790324 | rs775577612 | A | G |  |  | - | 3.25E-05 | 30746 | 25.5 | 0.912 | deleterious | possibly_damaging | Uncertain significance |
| *APOA5* | 11 | 116790340 | rs978653667 | T | C |  |  | - | 1.30E-04 | 30746 | 23.6 | 0.999 | tolerated | probably_damaging | Uncertain significance |
| *APOA5* | 11 | 116790346 | rs765011895 | A | G |  | S3, SD | - | 3.25E-05 | 30746 | 39 | 1 | NA | NA | not in ClinVar |
| *APOA5* | 11 | 116790355 | rs766805532 | G | T |  |  | - | 3.25E-05 | 30744 | 25.8 | 0.97 | deleterious | probably_damaging | Uncertain significance |
| *APOA5* | 11 | 116790357 | rs1270779185 | A | T |  |  | - | 6.51E-05 | 30746 | 25.9 | 1 | deleterious | probably_damaging | Uncertain significance |
| *APOA5* | 11 | 116790385 | rs778114184 | A | G |  |  | - | 3.25E-05 | 30746 | 24.7 | 0.863 | deleterious | possibly_damaging | not in ClinVar |
| *APOA5* | 11 | 116790389 | NA | G | C |  |  |  | 3.25E-05 | 30746 | 20.3 | 0.526 | deleterious | possibly_damaging | not in ClinVar |
| *APOA5* | 11 | 116790406 | rs149808404 | A | G |  |  | - | 3.25E-05 | 30746 | 35 | 1 | NA | NA | Conflicting classifications of pathogenicity |
| *APOA5* | 11 | 116790462 | NA | A | T |  |  |  | 3.25E-05 | 30746 | 23.3 | 0.721 | deleterious | possibly_damaging | not in ClinVar |
| *APOA5* | 11 | 116790463 | rs927429381 | A | C |  |  | - | 3.25E-05 | 30746 | 36 | 1 | NA | NA | not in ClinVar |
| *APOA5* | 11 | 116790489 | rs774692220 | C | T |  |  | - | 1.63E-04 | 30746 | 24.6 | 0.822 | deleterious | possibly_damaging | Uncertain significance |
| *APOA5* | 11 | 116790496 | rs376196775 | A | G |  |  | - | 9.76E-05 | 30746 | 22.1 | 1 | deleterious | possibly_damaging | Uncertain significance |
| *APOA5* | 11 | 116790501 | rs1940981051 | G | T |  |  |  | 3.25E-05 | 30746 | 25 | 0.986 | deleterious | probably_damaging | not in ClinVar |
| *APOA5* | 11 | 116790514 | NA | T | C |  |  |  | 3.25E-05 | 30746 | 25.2 | 0.871 | deleterious | probably_damaging | not in ClinVar |
| *APOA5* | 11 | 116790550 | rs748861443 | G | A |  |  | - | 3.25E-05 | 30746 | 23.5 | 1 | tolerated | possibly_damaging | not in ClinVar |
| *APOA5* | 11 | 116790565 | rs772514533 | T | C |  |  | - | 6.51E-05 | 30746 | 13.1 | 1 | tolerated | benign | not in ClinVar |
| *APOA5* | 11 | 116790585 | rs76753536 | A | G | 1 |  | - | 2.86E-03 | 30746 | 25.2 | 0.994 | deleterious | probably_damaging | Conflicting classifications of pathogenicity |
| *APOA5* | 11 | 116790616 | rs753285841 | A | G |  |  | - | 3.25E-05 | 30746 | 24.3 | 0.882 | deleterious | possibly_damaging | not in ClinVar |
| *APOA5* | 11 | 116790622 | rs778493133 | G | C |  |  | - | 9.76E-05 | 30746 | 21.8 | 0.718 | tolerated | benign | Uncertain significance |
| *APOA5* | 11 | 116790630 | rs780433260 | T | C |  |  | - | 9.76E-05 | 30746 | 14.51 | 1 | deleterious | benign | not in ClinVar |
| *APOA5* | 11 | 116790676 | rs2075291 | A | C | 1 | Mild, SD | 2.30E-03 | 2.86E-03 | 30746 | 20.3 | 1 | deleterious | probably_damaging | Conflicting classifications of pathogenicity |
| *APOA5* | 11 | 116790795 | rs368739905 | C | T | 1 |  | - | 6.51E-05 | 30744 | 14.37 | 1 | tolerated | benign | Uncertain significance |
| *APOA5* | 11 | 116790828 | NA | C | A |  |  |  | 3.25E-05 | 30740 | 10.66 | 1 | tolerated | benign | not in ClinVar |
| *APOA5* | 11 | 116790892 | NA | T | C |  |  |  | 3.25E-05 | 30746 | 22.4 | 0.999 | deleterious | benign | not in ClinVar |
| *APOA5* | 11 | 116790897 | rs1940997009 | T | A |  |  |  | 3.25E-05 | 30746 | 23.1 | 0.883 | deleterious | benign | Uncertain significance |
| *APOA5* | 11 | 116790910 | rs1940997395 | A | G |  |  |  | 3.25E-05 | 30746 | 24.6 | 0.569 | deleterious | possibly_damaging | not in ClinVar |
| *APOA5* | 11 | 116790912 | rs1434092204 | T | C |  |  | - | 9.76E-05 | 30746 | 23.6 | 0.963 | deleterious | benign | Uncertain significance |
| *APOA5* | 11 | 116790916 | rs146964666 | A | C |  |  | - | 1.95E-04 | 30746 | 13.87 | 1 | tolerated | benign | Uncertain significance |
| *APOA5* | 11 | 116790949 | rs201686049 | A | G |  |  | - | 3.25E-05 | 30746 | 23.5 | 1 | deleterious | benign | Uncertain significance |
| *APOA5* | 11 | 116790951 | rs757827003 | T | C |  |  | - | 3.25E-05 | 30746 | 17.77 | 1 | tolerated | benign | not in ClinVar |
| *APOA5* | 11 | 116790952 | rs867592586 | A | G |  |  | - | 3.25E-05 | 30744 | 20.7 | 1 | deleterious | benign | not in ClinVar |
| *APOA5* | 11 | 116790954 | NA | G | A |  |  |  | 3.25E-05 | 30746 | 19.08 | 1 | deleterious | benign | not in ClinVar |
| *APOA5* | 11 | 116791005 | rs773108378 | T | C |  |  | - | 3.25E-05 | 30746 | 13.5 | 1 | tolerated | benign | not in ClinVar |
| *APOA5* | 11 | 116791030 | rs372084940 | C | T |  |  | - | 3.25E-05 | 30744 | 12.47 | 1 | tolerated | benign | not in ClinVar |
| *APOA5* | 11 | 116791617 | rs1941014313 | A | T |  |  |  | 9.76E-05 | 30744 | 14.67 | 0.997 | tolerated | benign | not in ClinVar |
| *APOA5* | 11 | 116791623 | NA | G | C |  |  |  | 3.25E-05 | 30746 | 21.1 | 0.995 | deleterious | benign | not in ClinVar |
| *APOA5* | 11 | 116791636 | rs34282181 | T | G | 1 | SD | 3.86E-02 | 0.03728* | 30744 | 13.43 | 0.999 | tolerated | benign | Benign/Likely benign |
| *APOA5* | 11 | 116791658 | rs1941015800 | A | T |  |  |  | 3.25E-05 | 30746 | 24.3 | 0.741 | deleterious | possibly_damaging | not in ClinVar |
| *APOA5* | 11 | 116791683 | rs751538202 | A | G |  |  | - | 6.51E-05 | 30746 | 35 | 1 | NA | NA | not in ClinVar |
| *APOA5* | 11 | 116791691 | rs3135506 | A | G |  |  | - | 2.28E-04 | 30746 | 19.86 | 1 | deleterious | benign | Benign/Likely benign |
| *APOA5* | 11 | 116791691 | rs3135506 | C | G | 1 | SD | 6.73E-02 | 0.0629* | 30746 | 23.2 | 1 | deleterious | possibly_damaging | Benign/Likely benign |
| *APOA5* | 11 | 116791811 | NA | A | C |  |  |  | 3.25E-05 | 30744 | 25 | 1 | NA | NA | not in ClinVar |
| *LMF1* | 16 | 854548 | rs199591347 | C | A |  | SD | 0 | 3.25E-05 | 30744 | 12.97 | 1 | tolerated | benign | Likely benign |
| *LMF1* | 16 | 854551 | rs4984948 | C | G | 1 | SD | 7.60E-03 | 8.00E-03 | 30746 | 23.3 | 1 | deleterious | benign | Benign |
| *LMF1* | 16 | 854561 | rs199544373 | A | G |  | S4, SD | 0 | 6.51E-05 | 30744 | 23.2 | 1 | deleterious | probably_damaging | Uncertain significance |
| *LMF1* | 16 | 854599 | rs199843015 | A | G |  |  | - | 3.25E-05 | 30746 | 25.3 | 1 | deleterious | possibly_damaging | Uncertain significance |
| *LMF1* | 16 | 854615 | rs377058908 | T | C |  |  | 0 | 3.25E-05 | 30746 | 24.4 | 1 | deleterious | probably_damaging | not in ClinVar |
| *LMF1* | 16 | 854626 | rs533542876 | T | C |  | SD | 8.00E-04 | 1.63E-04 | 30746 | 26.8 | 0.998 | deleterious | probably_damaging | Uncertain significance |
| *LMF1* | 16 | 854668 | rs151137164 | T | C | 1 | SD | 3.40E-02 | 0.0267* | 30744 | 16.41 | 0.924 | deleterious | benign | Benign |
| *LMF1* | 16 | 854669 | rs758116895 | T | G |  |  | - | 6.51E-05 | 30746 | 22.8 | 0.92 | tolerated | benign | Uncertain significance |
| *LMF1* | 16 | 854698 | rs748287562 | T | C |  |  | - | 9.76E-05 | 30746 | 29.1 | 0.997 | deleterious | probably_damaging | Uncertain significance |
| *LMF1* | 16 | 854699 | rs772298418 | C | G |  |  |  | 3.25E-05 | 30746 | 26.7 | 0.985 | deleterious | probably_damaging | not in ClinVar |
| *LMF1* | 16 | 868948 | rs989431710 | T | G |  |  | - | 3.25E-05 | 30746 | 24.7 | 1 | deleterious | probably_damaging | Uncertain significance |
| *LMF1* | 16 | 868950 | rs372213215 | A | G |  |  | - | 1.30E-04 | 30746 | 24.1 | 0.94 | deleterious | probably_damaging | not in ClinVar |
| *LMF1* | 16 | 868973 | rs2069692739 | T | G |  |  |  | 3.25E-05 | 30746 | 12.46 | 1 | tolerated | benign | not in ClinVar |
| *LMF1* | 16 | 868992 | rs376340194 | C | G | 1 |  | - | 6.83E-04 | 30746 | 22.4 | 0.998 | deleterious | possibly_damaging | Uncertain significance |
| *LMF1* | 16 | 869002 | rs532127028 | T | C |  | SD | 0 | 3.25E-05 | 30746 | 23.6 | 1 | deleterious | possibly_damaging | Conflicting classifications of pathogenicity |
| *LMF1* | 16 | 869022 | rs1181212853 | A | G |  |  | - | 6.51E-05 | 30746 | 20.6 | 0.995 | tolerated | benign | Uncertain significance |
| *LMF1* | 16 | 869030 | rs372490098 | C | G |  |  | - | 6.51E-05 | 30746 | 15.84 | 0.737 | deleterious | benign | Likely benign |
| *LMF1* | 16 | 869041 | rs377384616 | A | C |  |  | - | 9.76E-05 | 30746 | 23.7 | 0.504 | deleterious | benign | Uncertain significance |
| *LMF1* | 16 | 869048 | rs770446199 | G | C | 1 |  | - | 9.76E-05 | 30746 | 22.5 | 0.991 | tolerated | possibly_damaging | not in ClinVar |
| *LMF1* | 16 | 869893 | rs574656841 | A | G |  | SD | 0 | 1.95E-04 | 30746 | 25.8 | 1 | deleterious | probably_damaging | Uncertain significance |
| *LMF1* | 16 | 869894 | rs181731943 | T | C |  | S2, S3, S4, Mild, SD | 0 | 2.60E-04 | 30746 | 24.7 | 1 | deleterious | probably_damaging | Conflicting classifications of pathogenicity |
| *LMF1* | 16 | 869903 | rs767509063 | A | T |  |  | - | 3.25E-05 | 30742 | 21.4 | 1 | tolerated | benign | Uncertain significance |
| *LMF1* | 16 | 869909 | rs1160512664 | G | A |  |  | - | 6.51E-05 | 30744 | 29.9 | 1 | deleterious | probably_damaging | not in ClinVar |
| *LMF1* | 16 | 869929 | rs757628838 | A | G |  |  | - | 3.25E-05 | 30746 | 32 | 1 | deleterious | probably_damaging | not in ClinVar |
| *LMF1* | 16 | 869948 | rs138205062 | A | G | 1 | S2, S3, S4, SD | 8.00E-04 | 5.20E-04 | 30746 | 25.3 | 1 | deleterious | probably_damaging | Benign/Likely benign |
| *LMF1* | 16 | 869981 | rs778629426 | T | C |  |  | - | 3.25E-05 | 30746 | 25.2 | 0.982 | deleterious | possibly_damaging | Uncertain significance |
| *LMF1* | 16 | 870002 | rs201927375 | T | C |  | SD | 0 | 1.95E-04 | 30746 | 12.89 | 1 | tolerated | benign | Uncertain significance |
| *LMF1* | 16 | 870007 | rs115416993 | T | G | 1 |  | 2.42E-02 | 0.02713* | 30744 | 11.51 | 1 | tolerated | benign | Benign/Likely benign |
| *LMF1* | 16 | 870028 | rs370807438 | C | G |  |  | - | 1.95E-04 | 30746 | 26 | 1 | deleterious | probably_damaging | Uncertain significance |
| *LMF1* | 16 | 870733 | rs199713950 | T | C |  | S4, SD | 0 | 3.25E-05 | 30746 | 23.5 | 1 | deleterious | probably_damaging | Likely benign |
| *LMF1* | 16 | 870735 | rs765778525 | C | A |  |  | - | 1.95E-04 | 30746 | 26.1 | 1 | deleterious | probably_damaging | not in ClinVar |
| *LMF1* | 16 | 870742 | rs376753256 | T | C |  |  | - | 6.51E-05 | 30746 | 25.1 | 1 | deleterious | probably_damaging | Uncertain significance |
| *LMF1* | 16 | 870753 | rs757459816 | G | A |  |  | - | 3.25E-05 | 30746 | 24.9 | 1 | deleterious | possibly_damaging | Uncertain significance |
| *LMF1* | 16 | 870775 | rs368805282 | A | G |  |  | - | 3.90E-04 | 30746 | 19.05 | 0.999 | deleterious | benign | not in ClinVar |
| *LMF1* | 16 | 870777 | rs186694298 | C | G |  |  | - | 1.63E-04 | 30746 | 16.77 | 0.841 | tolerated | benign | Conflicting classifications of pathogenicity |
| *LMF1* | 16 | 870789 | rs763603573 | C | T |  |  | - | 6.51E-05 | 30746 | 27.2 | 1 | deleterious | probably_damaging | not in ClinVar |
| *LMF1* | 16 | 870852 | rs769234511 | A | G |  |  | - | 9.76E-05 | 30746 | 14.86 | 1 | tolerated | benign | not in ClinVar |
| *LMF1* | 16 | 870858 | rs760931691 | A | T |  |  | - | 6.51E-05 | 30742 | 22.9 | 1 | deleterious | benign | not in ClinVar |
| *LMF1* | 16 | 870862 | rs936184702 | T | C |  |  | - | 3.25E-05 | 30746 | 16.79 | 1 | tolerated | benign | Uncertain significance |
| *LMF1* | 16 | 870870 | rs35168378 | T | C | 1 | SD | 4.84E-02 | 0.03506* | 30746 | 24.4 | 1 | deleterious | possibly_damaging | Benign |
| *LMF1* | 16 | 870871 | rs540153402 | A | G |  |  | - | 3.25E-05 | 30746 | 26 | 1 | deleterious | probably_damaging | Uncertain significance |
| *LMF1* | 16 | 871179 | rs143076454 | A | G | 1 |  | 3.00E-03 | 4.39E-03 | 30746 | 10.15 | 1 | tolerated | benign | Benign |
| *LMF1* | 16 | 871215 | rs776584760 | A | G |  |  | - | 1.30E-04 | 30746 | 37 | 1 | NA | NA | Conflicting classifications of pathogenicity |
| *LMF1* | 16 | 871281 | rs1439916071 | C | G |  |  | - | 3.25E-05 | 30742 | 12.39 | 0.63 | tolerated | benign | not in ClinVar |
| *LMF1* | 16 | 871282 | rs774248480 | C | G |  |  |  | 3.25E-05 | 30744 | 24.4 | 0.998 | deleterious | probably_damaging | Likely benign |
| *LMF1* | 16 | 871301 | rs753503972 | A | C |  |  | - | 3.25E-05 | 30738 | 24.7 | 1 | deleterious | benign | Uncertain significance |
| *LMF1* | 16 | 871323 | rs372159961 | T | C |  | Mild | - | 3.25E-05 | 30746 | 25.5 | 1 | deleterious | probably_damaging | not in ClinVar |
| *LMF1* | 16 | 879571 | rs577358020 | C | T |  |  | 8.00E-04 | 3.25E-05 | 30744 | 34 | 1 | deleterious | probably_damaging | Likely pathogenic |
| *LMF1* | 16 | 879613 | rs374533571 | T | C |  |  | - | 2.28E-04 | 30746 | 24.4 | 0.721 | tolerated | probably_damaging | Uncertain significance |
| *LMF1* | 16 | 879629 | rs372333537 | G | A |  |  | - | 1.30E-04 | 30724 | 22.7 | 0.998 | tolerated | benign | Likely benign |
| *LMF1* | 16 | 879630 | rs61745065 | T | G | 1 | SD | 9.83E-02 | 0.08869* | 30746 | 23.8 | 0.996 | tolerated | benign | Benign |
| *LMF1* | 16 | 879647 | rs531415593 | T | C |  | SD | 0 | 3.25E-05 | 30746 | 28 | 1 | deleterious | probably_damaging | Uncertain significance |
| *LMF1* | 16 | 879667 | rs754428234 | A | G |  |  | - | 6.51E-05 | 30746 | 24.6 | 0.993 | deleterious | probably_damaging | Conflicting classifications of pathogenicity |
| *LMF1* | 16 | 879700 | rs1364874340 | C | T |  |  | - | 3.25E-05 | 30746 | 24.6 | 1 | deleterious | probably_damaging | not in ClinVar |
| *LMF1* | 16 | 879730 | rs1307363051 | A | G |  |  |  | 1.30E-04 | 30746 | 27.2 | 1 | deleterious | probably_damaging | not in ClinVar |
| *LMF1* | 16 | 893006 | NA | T | C |  |  |  | 3.25E-05 | 30746 | 24.9 | 1 | NA | NA | not in ClinVar |
| *LMF1* | 16 | 893011 | rs754165819 | C | T |  |  | - | 1.30E-04 | 30730 | 25.8 | 1 | deleterious | probably_damaging | Uncertain significance |
| *LMF1* | 16 | 893018 | rs34295987 | G | A | 1 | SD | 2.30E-03 | 3.35E-03 | 30738 | 24.8 | 1 | deleterious | benign | Conflicting classifications of pathogenicity |
| *LMF1* | 16 | 893022 | rs1475433901 | G | C |  |  | - | 3.25E-05 | 30746 | 24.9 | 1 | deleterious | possibly_damaging | not in ClinVar |
| *LMF1* | 16 | 893023 | rs370864352 | G | A |  | S2, S3, SD | - | 3.25E-05 | 30746 | 25.4 | 1 | deleterious | probably_damaging | not in ClinVar |
| *LMF1* | 16 | 893033 | rs1266228331 | A | G |  |  | - | 3.25E-05 | 30746 | 24.7 | 0.998 | deleterious | probably_damaging | not in ClinVar |
| *LMF1* | 16 | 893035 | rs1424071356 | C | T |  |  | - | 3.25E-05 | 30744 | 23.6 | 0.998 | deleterious | benign | not in ClinVar |
| *LMF1* | 16 | 893039 | rs199953320 | A | G |  |  | - | 3.25E-05 | 30746 | 38 | 1 | NA | NA | Likely pathogenic |
| *LMF1* | 16 | 893048 | rs376563644 | A | G |  |  | 8.00E-04 | 4.88E-04 | 30746 | 24.5 | 1 | deleterious | possibly_damaging | Uncertain significance |
| *LMF1* | 16 | 893051 | rs1367017632 | T | C |  |  | - | 3.25E-05 | 30746 | 24.5 | 1 | deleterious | probably_damaging | not in ClinVar |
| *LMF1* | 16 | 893053 | rs754772870 | A | C |  |  | - | 3.25E-05 | 30746 | 24.1 | 1 | deleterious | probably_damaging | Uncertain significance |
| *LMF1* | 16 | 893053 | rs754772870 | T | C |  | S3 | - | 6.51E-05 | 30746 | 24 | 0.987 | deleterious | possibly_damaging | Uncertain significance |
| *LMF1* | 16 | 893057 | rs573828508 | A | G |  |  | 0 | 6.51E-05 | 30746 | 27.4 | 0.997 | deleterious | probably_damaging | Uncertain significance |
| *LMF1* | 16 | 893072 | NA | T | C |  |  |  | 6.51E-05 | 30746 | 26.8 | 1 | deleterious | probably_damaging | not in ClinVar |
| *LMF1* | 16 | 910962 | rs182946890 | T | C |  | SD | 0 | 3.25E-05 | 30746 | 25.2 | 0.599 | deleterious | possibly_damaging | not in ClinVar |
| *LMF1* | 16 | 910984 | rs375836686 | A | G |  |  | - | 3.25E-05 | 30744 | 13.83 | 0.879 | tolerated | benign | Uncertain significance |
| *LMF1* | 16 | 910989 | NA | A | G |  |  |  | 3.25E-05 | 30746 | 20.7 | 1 | tolerated | benign | not in ClinVar |
| *LMF1* | 16 | 911019 | rs370179235 | A | G |  |  | - | 3.90E-04 | 30746 | 23 | 0.764 | deleterious | possibly_damaging | Uncertain significance |
| *LMF1* | 16 | 934244 | rs201406396 | T | C |  | SD | 0 | 6.51E-05 | 30746 | 24.4 | 0.702 | deleterious | probably_damaging | Likely pathogenic |
| *LMF1* | 16 | 934246 | rs376658034 | G | A |  |  | - | 3.25E-05 | 30746 | 25.3 | 0.676 | deleterious | probably_damaging | Uncertain significance |
| *LMF1* | 16 | 934249 | rs1233828390 | A | G |  |  | - | 6.51E-05 | 30746 | 24.6 | 0.829 | deleterious | probably_damaging | not in ClinVar |
| *LMF1* | 16 | 954369 | rs35663121 | G | A | 1 | SD | 8.55E-02 | 0.07474* | 30746 | 22.2 | 0.999 | tolerated | benign | Benign |
| *LMF1* | 16 | 954394 | rs763405270 | T | C |  |  | - | 2.60E-04 | 30746 | 22.1 | 1 | deleterious | benign | not in ClinVar |
| *LMF1* | 16 | 954412 | rs922113767 | C | G | 1 |  | - | 9.76E-05 | 30746 | 20.5 | 0.898 | tolerated | benign | Uncertain significance |
| *LMF1* | 16 | 954426 | rs771658591 | T | C |  |  | - | 1.30E-04 | 30746 | 17.46 | 0.994 | tolerated | benign | Uncertain significance |
| *LMF1* | 16 | 954430 | NA | T | C |  |  |  | 3.25E-05 | 30746 | 22.9 | 0.996 | deleterious | benign | Uncertain significance |
| *LMF1* | 16 | 954432 | rs375529211 | A | G |  | SD | 0 | 3.25E-05 | 30746 | 15.48 | 0.877 | tolerated | benign | Uncertain significance |
| *LMF1* | 16 | 954463 | rs748153798 | T | C |  |  | - | 6.51E-05 | 30742 | 24.2 | 1 | deleterious | probably_damaging | not in ClinVar |
| *LMF1* | 16 | 954466 | rs1030852813 | A | G |  |  | - | 3.25E-05 | 30746 | 15.53 | 1 | tolerated | possibly_damaging | not in ClinVar |
| *LMF1* | 16 | 954487 | rs1323912190 | G | T |  |  | - | 3.25E-05 | 30746 | 15.87 | 1 | tolerated | benign | Uncertain significance |
| *LMF1* | 16 | 954497 | rs1274682128 | C | G |  |  | - | 3.25E-05 | 30744 | 13.96 | 0.982 | tolerated | benign | Uncertain significance |
| *LMF1* | 16 | 954507 | rs1047361482 | C | T |  |  | - | 9.76E-05 | 30742 | 22.8 | 1 | tolerated | benign | not in ClinVar |
| *LMF1* | 16 | 954536 | rs1230523425 | C | G |  |  | - | 3.25E-05 | 30746 | 16.2 | 1 | tolerated | benign | not in ClinVar |
| *LMF1* | 16 | 954562 | rs35124265 | T | C |  | SD | - | 3.25E-05 | 30746 | 19.13 | 1 | tolerated | benign | Uncertain significance |
| *LMF1* | 16 | 954622 | rs1045081351 | T | C |  |  | - | 3.25E-05 | 30746 | 24 | 1 | deleterious | possibly_damaging | not in ClinVar |
| *LMF1* | 16 | 954625 | rs372696701 | G | T | 1 |  | 1.50E-03 | 1.40E-03 | 30746 | 21.9 | 0.999 | tolerated | benign | not in ClinVar |
| *LMF1* | 16 | 954661 | rs779735217 | G | C |  |  | - | 6.51E-05 | 30746 | 26.5 | 1 | deleterious | probably_damaging | Uncertain significance |
| *LMF1* | 16 | 954664 | rs753472646 | T | C |  |  | - | 3.25E-05 | 30746 | 25.5 | 1 | deleterious | possibly_damaging | Uncertain significance |
| *LMF1* | 16 | 970793 | rs370716130 | T | A |  |  | - | 9.76E-05 | 30742 | 25 | 0.682 | deleterious | possibly_damaging | Uncertain significance |
| *LMF1* | 16 | 970823 | rs1348395304 | T | C |  |  | - | 3.25E-05 | 30746 | 25.8 | 1 | deleterious | probably_damaging | not in ClinVar |
| *LMF1* | 16 | 970842 | NA | A | C |  |  |  | 3.25E-05 | 30746 | 25.1 | 0.999 | deleterious | probably_damaging | not in ClinVar |
| *LMF1* | 16 | 970844 | rs974624219 | A | G |  |  | - | 1.95E-04 | 30746 | 17.83 | 1 | deleterious | benign | not in ClinVar |
| *LMF1* | 16 | 970868 | rs772359935 | A | G |  |  | - | 3.25E-05 | 30746 | 11.55 | 1 | tolerated | benign | Uncertain significance |
| *LMF1* | 16 | 970877 | rs759097865 | A | C |  |  | - | 3.25E-05 | 30746 | 11.71 | 1 | tolerated | benign | not in ClinVar |
| *LMF1* | 16 | 970877 | rs759097865 | G | C |  |  | - | 3.25E-05 | 30746 | 12.23 | 1 | tolerated | benign | not in ClinVar |
| *LMF1* | 16 | 970889 | rs368337185 | A | G |  |  | - | 8.46E-04 | 30746 | 19.79 | 1 | deleterious | benign | Uncertain significance |
| *LMF1* | 16 | 970940 | NA | A | G |  |  |  | 3.25E-05 | 30746 | 17.36 | 1 | tolerated | benign | Uncertain significance |
| *LMF1* | 16 | 970979 | rs1327180945 | G | A |  |  |  | 3.26E-05 | 30692 | 17.35 | 0.991 | deleterious | benign | not in ClinVar |
| *APOC2* | 19 | 44948525 | NA | C | T |  |  |  | 3.25E-05 | 30744 | 24.9 | 1 | deleterious | possibly_damaging | not in ClinVar |
| *APOC2* | 19 | 44948530 | rs1970347563 | G | T |  |  |  | 3.26E-05 | 30726 | 12.86 | 1 | tolerated | benign | Uncertain significance |
| *APOC2* | 19 | 44948703 | rs201709243 | A | G |  |  | - | 4.23E-04 | 30744 | 17.57 | 1 | tolerated | benign | not in ClinVar |
| *APOC2* | 19 | 44948730 | rs147242592 | A | G | 1 |  | 2.30E-03 | 2.63E-03 | 30746 | 22.5 | 0.958 | deleterious | probably_damaging | Conflicting classifications of pathogenicity |
| *APOC2* | 19 | 44948746 | rs200404502 | T | C |  |  | 0 | 9.76E-05 | 30746 | 10.28 | 1 | tolerated | benign | Uncertain significance |
| *APOC2* | 19 | 44948767 | rs120074114 | C | A |  | S2, S3, S4, SD | 0 | 9.76E-05 | 30742 | 15.92 | 0 | deleterious | benign | Conflicting classifications of pathogenicity |
| *APOC2* | 19 | 44948776 | NA | C | T |  |  |  | 3.25E-05 | 30744 | 23.2 | 0.948 | deleterious | probably_damaging | not in ClinVar |
| *APOC2* | 19 | 44948806 | rs761724352 | G | C |  |  | - | 3.25E-05 | 30746 | 20.2 | 1 | deleterious | possibly_damaging | not in ClinVar |
| *APOC2* | 19 | 44948808 | rs750370010 | A | G |  |  | - | 6.51E-05 | 30746 | 19.73 | 1 | tolerated | probably_damaging | Uncertain significance |
| *APOC2* | 19 | 44948850 | rs148445956 | A | G |  |  | - | 6.51E-05 | 30746 | 24.8 | 1 | deleterious | probably_damaging | Uncertain significance |
| *APOC2* | 19 | 44949166 | rs1339593526 | C | T |  |  | - | 6.51E-05 | 30716 | 26.6 | 1 | deleterious | probably_damaging | not in ClinVar |
| *APOC2* | 19 | 44949189 | rs1324823081 | A | G |  |  | - | 3.25E-05 | 30732 | 16.92 | 1 | tolerated | benign | not in ClinVar |
| *APOC2* | 19 | 44949200 | rs199687805 | G | C | 1 |  | - | 3.25E-05 | 30746 | 17.57 | 1 | tolerated | possibly_damaging | Uncertain significance |

*Common variant (MAF >0.01); ^†^Carriage by at least one individual in the severe HTG or mild-to-moderate HTG groups

LPL = Lipoprotein Lipase; GPIHBP1 = Glycosylphosphatidylinositol Anchored High Density Lipoprotein Binding Protein 1; APOA5 = Apolipoprotein A5; LMF1 = Lipase Maturation Factor 1; APOC2 = Apolipoprotein C2; MAF = minor allele frequency

S2=Dron et al, Severe hypertriglyceridemia is primarily polygenic. *J Clin Lipidol*. 2019, Table S2; S3=Dron et al, Severe hypertriglyceridemia is primarily polygenic. *J Clin Lipidol*. 2019, Table S3; S4=Dron et al, Severe hypertriglyceridemia is primarily polygenic. *J Clin Lipidol*. 2019, Table S4; Mild= The polygenic nature of mild-to-moderate hypertriglyceridemia. *J Clin Lipidol*. 2020, Table S2; SD=Gill et al, Ancestry-specific profiles of genetic determinants of severe hypertriglyceridemia. *J Clin Lipidol*. 2021, Table S2.

**Supplemental Table S6.** Accumulation of Genetic Risk Factors in the Mild-to-moderate HTG and Severe HTG categories, Compared to Normal TG Controls—Primary Analyses.

| TG categories | Genetic Risk Factors | N Carriers | N Non-carriers | Odds Ratio | 95% CI | P-value |
| --- | --- | --- | --- | --- | --- | --- |
| Normal TG (Reference) | Carriers of rare variants | 383 | 3149 | Ref | Ref | Ref |
|  | APOA5 p.S19W carriage | 467 | 3065 | Ref | Ref | Ref |
|  | Top 10% PRS | 354 | 3178 | Ref | Ref | Ref |
|  | Top 10% potentially causal TG variants | 382 | 3150 | Ref | Ref | Ref |
|  | Any genetic risk factor | 1284 | 2248 | Ref | Ref | Ref |
| Mild-to-moderate HTG | Carriers of rare variants | 43 | 299 | 1.18 | 0.84, 1.66 | 0.33 |
|  | APOA5 p.S19W carriage | 78 | 264 | 1.94 | 1.48, 2.54 | 1.63E-06 |
|  | Top 10% PRS | 59 | 283 | 1.87 | 1.38, 2.53 | 4.54E-5 |
|  | Top 10% potentially causal TG variants | 59 | 283 | 1.72 | 1.27, 2.32 | 4.00E-4 |
|  | Any genetic risk factor | 174 | 168 | 1.81 | 1.45, 2.27 | 1.65E-07 |
| Severe HTG | Carriers of rare variants | ≤20 | ≤20 | 2.24 | 0.40, 8.54 | 0.19 |
|  | APOA5 p.S19W carriage | ≤20 | ≤20 | 3.65 | 1.22, 10.93 | 0.02 |
|  | Top 10% PRS | ≤20 | ≤20 | 0.69 | 0.02, 4.62 | 1.00 |
|  | Top 10% potentially causal TG variants | ≤20 | ≤20 | 2.25 | 0.40, 8.56 | 0.19 |
|  | Any genetic risk factor | ≤20 | ≤20 | 3.15 | 1.05, 9.42 | 0.04 |

HTG = Hypertriglyceridemia; TG = triglyceride; APOA5 = Apolipoprotein A5; PRS = Polygenic Risk Score; CI = confidence interval

**Supplemental Table S7. Association between Common Variants from 5 Canonical Triglyceride Gene Regions and Log-Transformed Median Measured Triglycerides Adjusted by Gender and Age.**

| **Chromosome** | **SNP** | **rsID** | **Gene in the region** | **A1** | **NMISS** | **Beta** | **Stat** | **P** |
| --- | --- | --- | --- | --- | --- | --- | --- | --- |
| 8 | chr8:143215394:C:T | rs78367243 | *GPIHBP1* | T | 15373 | -0.01465 | -1.969 | 0.04903 |
| 11 | chr11:116791636:G:T | rs34282181 | *APOA5* | T | 15372 | -0.05727 | -3.997 | 6.45E-05 |
| 11 | chr11:116791691:G:C | rs3135506 | *APOA5* | C | 15373 | 0.1165 | 10.49 | 1.15E-25 |
| 16 | chr16:854668:C:T | rs151137164 | *LMF1* | T | 15372 | 0.02882 | 1.722 | 0.08518 |
| 16 | chr16:870007:G:T | rs115416993 | *LMF1* | T | 15372 | 0.01033 | 0.6169 | 0.5373 |
| 16 | chr16:870870:C:T | rs35168378 | *LMF1* | T | 15373 | 0.007327 | 0.4956 | 0.6202 |
| 16 | chr16:879630:G:T | rs61745065 | *LMF1* | T | 15373 | 0.002744 | 0.2867 | 0.7743 |
| 16 | chr16:954369:A:G | rs35663121 | *LMF1* | G | 15373 | -0.007452 | -0.7193 | 0.472 |

**Supplemental Table S8. Polygenic Risk Scores for Triglycerides Derived from a Population with African Ancestry**(5, 6) **by Category.**

|  | Primary Analysis Categories | | | Sensitivity Analysis Categories | |
| --- | --- | --- | --- | --- | --- |
|  | **Severe HTG**  (>885 mg/dL) | **Mild-to-**  **moderate HTG**  (300-885 mg/dL) | **Normal TG** (79.40-149.40 mg/dL) | **AHA HTG**  (>500 mg/dL) | **Top 1% TG**  (>374.4 mg/dL) |
| N | ≤20 | 342 | 3532 | 54 | 154 |
| TG PRS, median [IQR] | -0.0003884  [-0.0006645, 0.0002023] | -0.0001513  [-0.0005752, 0.0003385] | -0.0003503  [-0.0008414, 0.0001109] | -0.0001052  [-0.0005353, 0.0003137] | -0.0001113  [-0.0006723, 0.0003422] |

PRS = polygenic risk score; HTG = high triglyceride; TG = triglyceride; AHA = American Heart Association; IQR = interquartile range

**Supplemental Table S9. Total Allele Count of Potentially Causal Triglyceride Variants** **by Category.**

|  | Primary Analysis Categories | | | Sensitivity Analysis Categories | |
| --- | --- | --- | --- | --- | --- |
|  | **Severe HTG**  (>885 mg/dL) | **Mild-to-**  **moderate HTG**  (300-885 mg/dL) | **Normal TG** (79.5-149 mg/dL) | **AHA HTG**  (>500 mg/dL) | **Top 1% TG**  (>374.4 mg/dL) |
| N | ≤20 | 342 | 3532 | 54 | 154 |
| Allele count, median [IQR] | 132.5  [130.2, 138.8] | 133  [130, 137] | 132  [129, 136] | 133.5  [131, 138] | 133  [130, 137] |

HTG = high triglyceride; TG = triglyceride; AHA = American Heart Association; IQR = interquartile range

**Supplemental Table S10. Cohort Characteristics—Sensitivity Analysis Categories (AHA Severe HTG and Top 1% TG).**

|  | | **AHA Severe HTG**  (>500 mg/dL) | **Top 1% TG**  (>374.4 mg/dL) |
| --- | --- | --- | --- |
| N | | 54 | 154 |
| Age (years), median [IQR] | | 55 [48-64] | 57 [49-64] |
| Gender, N (%) | Female | 29 (53.7) | 77 (50.0) |
|  | Male | 25 (46.3) | 77 (50.0) |
| BMI, median [IQR] | | 31.36 [27.03-36.06] | 32.58 [28.39-37.24] |
| Lipid-lowering medication, N (%) | | 42 (77.8) | 115 (74.7) |
| Adjusted triglycerides (mg/dL), median [IQR] | | 662.50 [550.20-874.8] | 460.60 [397.20-553.90] |

IQR=interquartile range; HTG = Hypertriglyceridemia; TG = triglyceride

**Supplemental Figure S1.** **Sensitivity Analyses: Frequencies of genetic factors in different TG categories (AHA Severe HTG, Mild-to-Moderate HTG, and Normal TG) in individuals of African ancestry.**

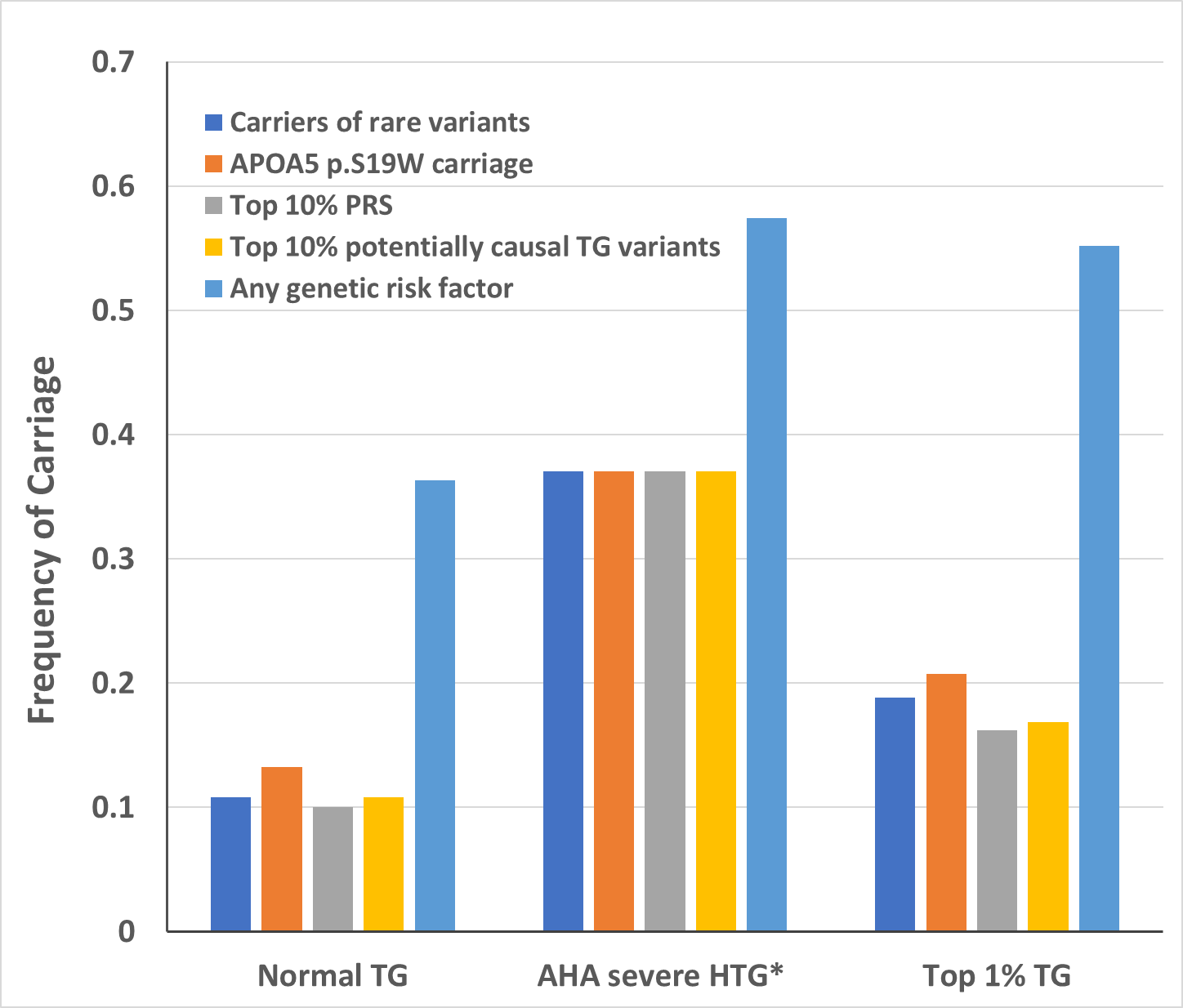

*Percentages are represented as ≤37.5% in accordance with AoU policy, which prohibits cells or percentages representing N<20.

HTG = Hypertriglyceridemia; TG = triglyceride; AHA = American Heart Association; APOA5 = Apolipoprotein A5; PRS = Polygenic Risk Score

**Supplemental Figure S2.** **Sensitivity Analyses: Forrest Plots for Genetic Risk Factors for Elevated TG Levels in Individuals of African ancestry. (AHA Severe HTG, Mild-to-Moderate HTG, and Normal TG).**

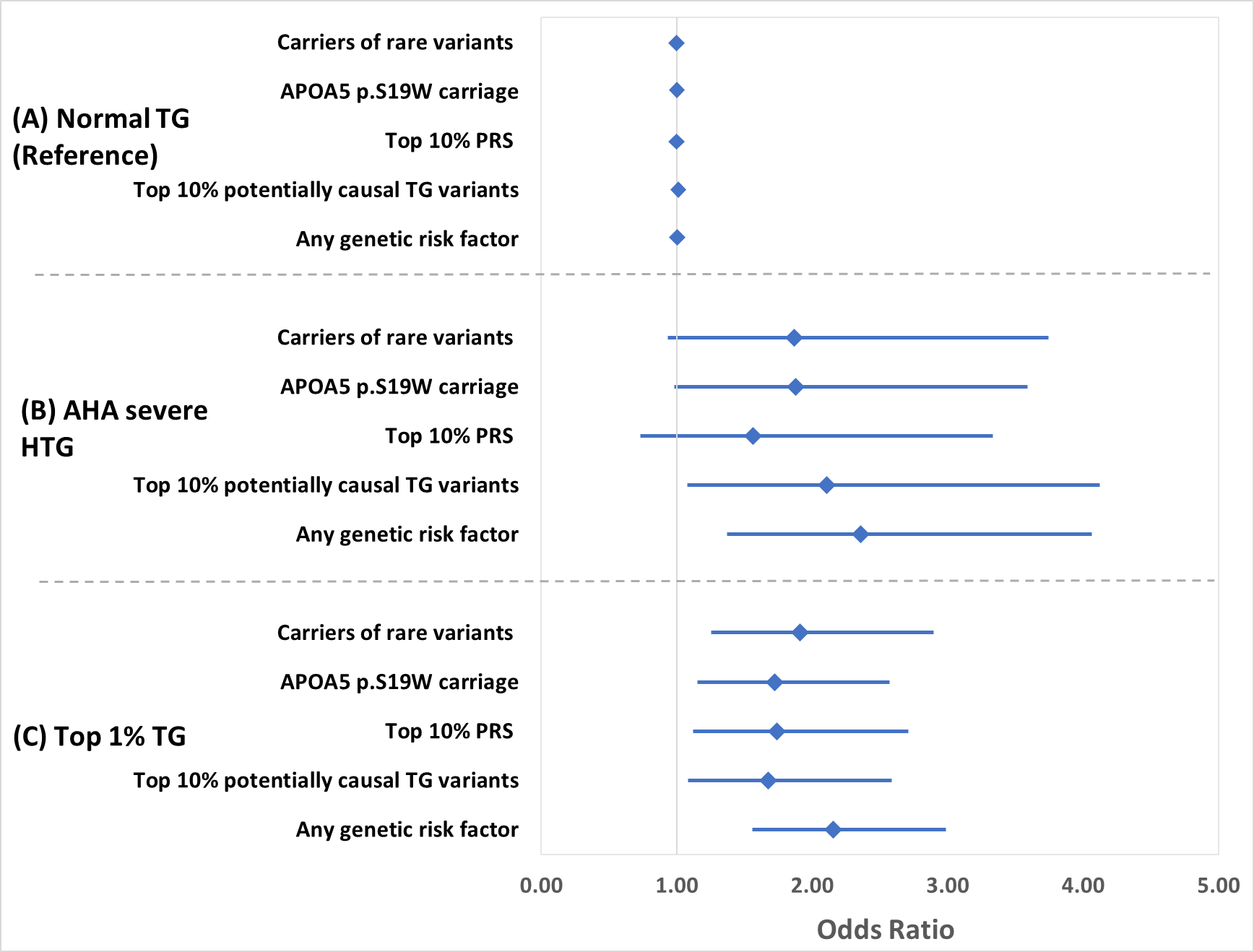

HTG = Hypertriglyceridemia; TG = triglyceride; AHA = American Heart Association; APOA5 = Apolipoprotein A5; PRS = Polygenic Risk Score

**Supplemental Table S11. Accumulation of Genetic Risk Factors in the AHA HTG, and Top 1% HTG categories, Compared to Normal TG Controls—Sensitivity Analyses. (finished editing)**

| TG categories | Genetic Risk Factors | N Carriers | N Non-carriers | Odds Ratio | 95% CI | P-value |
| --- | --- | --- | --- | --- | --- | --- |
| Normal TG (Reference) | Carriers of rare variants | 383 | 3149 | Ref | Ref | Ref |
|  | APOA5 p.S19W carriage | 467 | 3065 | Ref | Ref | Ref |
|  | Top 10% PRS | 354 | 3178 | Ref | Ref | Ref |
|  | Top 10% potentially causal TG variants | 382 | 3150 | Ref | Ref | Ref |
|  | Any genetic risk factor | 1284 | 2248 | Ref | Ref | Ref |
| AHA Severe HTG | Carriers of rare variants | ≤20* | >34* | 1.87 | 0.93, 3.74 | 0.078 |
|  | APOA5 p.S19W carriage | ≤20* | >34* | 1.88 | 0.98, 3.59 | 0.058 |
|  | Top 10% PRS | ≤20* | >34* | 1.56 | 0.73, 3.33 | 0.250 |
|  | Top 10% potentially causal TG variants | ≤20* | >34* | 2.11 | 1.08, 4.13 | 0.029 |
|  | Any genetic risk factor | 31 | 23 | 2.36 | 1.37, 4.06 | 0.002 |
| Top 1% HTG | Carriers of rare variants | 29 | 125 | 1.91 | 1.26, 2.90 | 0.002 |
|  | APOA5 p.S19W carriage | 32 | 122 | 1.72 | 1.15, 2.57 | 0.009 |
|  | Top 10% PRS | 25 | 129 | 1.74 | 1.12, 2.71 | 0.014 |
|  | Top 10% potentially causal TG variants | 26 | 128 | 1.67 | 1.08, 2.59 | 0.020 |
|  | Any genetic risk factor | 85 | 69 | 2.16 | 1.56, 2.98 | 3.54E-06 |

*Counts are presented as ≤20 and >34 for AHA Severe HTG (N=54) in accordance with AoU policy, which prohibits reporting cell counts or providing the ability to calculate N<20.

HTG = Hypertriglyceridemia; TG = triglyceride; APOA5 = Apolipoprotein A5; PRS = Polygenic Risk Score; AHA = American Heart Association; CI = confidence interval
